# Recovery from Heart Failure is a Vascular Recovery

**DOI:** 10.1101/2024.07.24.24310960

**Authors:** Rajul K. Ranka, Krishan Gupta, Felix Naegele, Alexander J. Lu, Shuang Li, Michael Graber, Kaylee N. Carter, Anahita Mojiri, Lili Zhang, Arvind Bhimaraj, Li Lai, Keith A. Youker, Kaifu Chen, John P. Cooke

**Author notes:** Corresponding Author: John P. Cooke, M.D., Ph.D. Joseph C. “Rusty” Walter and Carole Walter Looke Presidential Distinguished Chair Professor and Chair, Department of Cardiovascular Sciences Director, Center for Cardiovascular Regeneration Director, Center for RNA Therapeutics Chief Translational Officer, Houston Methodist Academic Institute President, Society for RNA Therapeutics 6670 Bertner Ave., Mail Stop: R10-South Houston, Texas 77030. Co-corresponding Author: Kaifu Chen, Ph.D. Associate Professor of Pediatrics, Harvard Medical School Director of Computational Biology Program Basic and Translational Research Division, Department of Cardiology, Boston Children’s Hospital, Boston, MA 02115.

## Abstract

Heart failure (HF) remains a major cause of morbidity and mortality worldwide, with limited treatment options. Heart transplantation is an end stage option but limited by donor availability. Left-ventricular assist device (LVAD) implantation serves as a bridging strategy for patients awaiting a transplant. Intriguingly, LVAD support (typically for 6-12 months before heart transplantation) is often associated with some level of improvement in cardiac function and histology. In rare cases, LVAD support can improve cardiac function sufficiently to avoid heart transplantation after LVAD removal. The underlying mechanisms of this improvement in cardiac function are not understood. Here, we provide evidence that the improvement in cardiac function post-LVAD is associated with a reduction in fibrosis and an increase in capillary density. This heart failure recovery (HFR) is also associated with an angiogenic cell fate transition. We observed a distinct pro-angiogenic phenotype of cardiac non-myocytes isolated from post-LVAD hearts. Single-nuclei RNA sequencing of pre- and post-LVAD cardiac tissue reveals a fibroblast subtype that undergoes mesenchymal to endothelial transition (MEndoT), potentially facilitating HFR. In a murine model of HFR, lineage tracing studies confirm that MEndoT is associated with the increase in capillary density and perfusion during HFR. In summary, our results support the new concept that HFR is associated with a reduction in interstitial cardiac fibrosis, an increase in capillary density and perfusion, that is due in part to an angiogenic cell fate transition. Our work represents a shift in the conceptual framework regarding mechanisms of HFR, and a new therapeutic avenue for exploration.

## Main

Heart failure (HF) is a major cause of morbidity and mortality, affecting more than 64 million people worldwide^1–3^. Myocardial infarction causes loss of functional cardiac muscle and is a major cause of HF, however 40% of HF cases are non-ischemic in nature i.e., HF in the absence of significant coronary artery disease^4, 5^. As opposed to the confluent scar observed after MI, in “non-ischemic” HF one observes disseminated interstitial and perivascular fibrosis which may be due in part to endothelial to mesenchymal transition^6^. The accumulation of extracellular matrix (ECM) components, particularly collagen, impairs cardiac function and is associated with adverse outcomes^7–9^.

Advances in medical therapy over the last two decades have improved patient outcomes in HF. Nevertheless, there is still a significant number of patients who progress to end-stage HF, necessitating advanced therapeutic strategies including heart transplantation^10^. Unfortunately, there is a limited availability of donor hearts with only 3,700 heart transplants performed each year. Therefore, the use of left ventricular assist devices (LVADs) has emerged as a therapeutic strategy, providing a bridge to recovery, remission, or transplantation for patients with advanced HF^11–13^.

LVAD support appears to promote favorable cellular and structural changes within the myocardium^14^. We and others have observed that the use of a LVAD device may be associated with improved ventricular compliance^15^, reduced cardiac hypertrophy^16, 17^, reduced cardiac fibrosis^16^, increased capillary density^18^ and bulk gene expression changes consistent with a mesenchyme to endothelial transition (MEndoT)^19^. These alterations may be associated with sufficient improvement in cardiac function that transplant is no longer necessary. This regenerative response to LVAD implantation is most associated with patients that have “non-ischemic” HF^20–24^.

To explore the mechanisms of HF recovery (HFR), we employed single nuclei transcriptomic analysis of pre- and post-LVAD hearts. In addition, we developed methods to culture non-myocytes (NMs) from these hearts for further phenotypic and functional characterization. Furthermore, we validated a murine model of HFR and combined it with a tamoxifen inducible fibroblast lineage tracing strategy to assess cell fate transitions in HFR. Our new work reveals that recovery from HF is in part a microvascular recovery. Specifically, angiogenic transdifferentiation expands the cardiac microvasculature to improve myocardial perfusion and is associated with favorable structural and functional changes of the heart. Our focus on molecular modulation of the myocardial microvasculature, and the role of angiogenic transdifferentiation^25–27^, has generated a novel conceptual framework to understand HFR, and to guide the next generation of HF therapies.

## Results

### Mechanical Unloading Reverses Hallmarks of Heart Failure

In this study we focused on patients with “non-ischemic” HF. Such patients typically have a history of diabetes and/or hypertension, and have impaired systolic and diastolic cardiac function, cardiac and myocyte hypertrophy associated with disseminated fibrosis and reduced capillary density^7–10^. We obtained LV myocardial samples from HF patients at the time of implantation of LVAD (pre-LVAD or HF group) and 6-18 months later at the time of LVAD explantation and heart transplant (post-LVAD or HFR group) (Fig.1a). Demographics of patients are presented in Supplemental Table 1. The patient cohort was composed of 16 patients (median age 53 years, 37.5% women) with end-stage HF, receiving LVAD implantation as bridge-to-transplantation therapy at Houston Methodist Hospital.

Masson’s trichrome stain was used to evaluate and quantify myocardial fibrosis in each sample from pre- or post-LVAD hearts (Fig. 1b). Tissue sections from post-LVAD hearts manifested less fibrosis than pre-LVAD hearts (10% vs. 19%, *p*=0.03, n=5); fewer fibroblasts (FBs) per low power field (LPF) (66.4±4 vs. 98.6±7, *p*<0.0001, n=5) (Fig. 1c, Supplemental Fig. 1a); and increased endothelial cells (ECs) per LPF (96.8±6 vs. 74.8±8, *p*<0.01, n=5) (Fig. 1c & 1e, Supplemental Fig. 1b). There was a negative correlation of FB count with EC count (*p*=0.01) (Fig. 1d).

**Figure 1.**
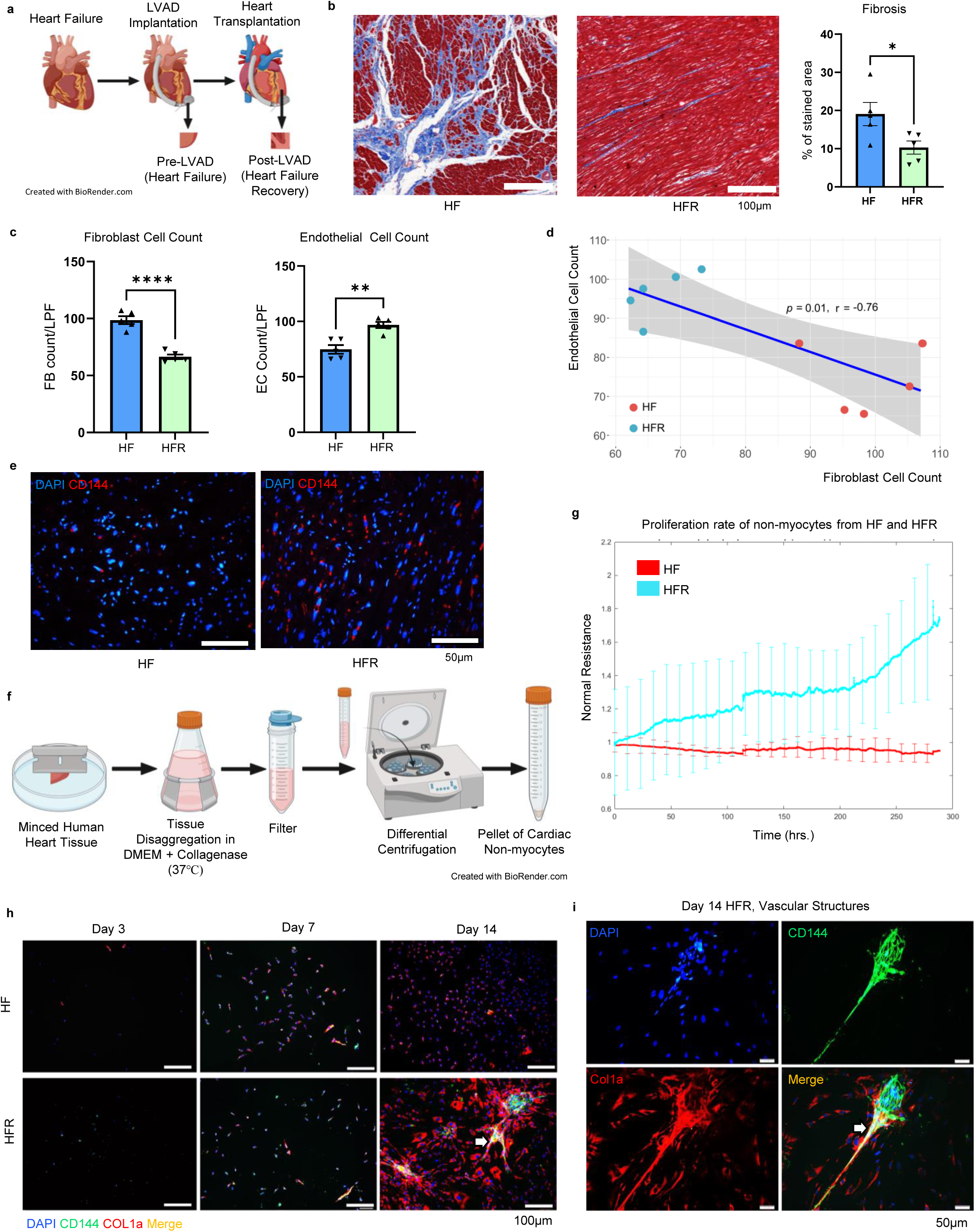
Human cardiac tissue and isolated cardiac non-myocytes from pre- and post-LVAD hearts indicate that a microvascular recovery contributes to HFR. **a,** Schematic of sample acquisition from pre-LVAD/HF and post-LVAD/HFR human hearts. **b,** Masson’s trichrome staining show reduction in fibrosis in HFR. **c,** FB cell count decreased whereas EC count increased in HFR hearts compared to HF. **d,** Pearson correlation analysis reveals a negative correlation between FB and EC count in HF and HFR tissue. **e,** CD144 staining shows increased number of ECs in HFR tissue. **f,** Schematic of isolation of human cardiac NMs from HF and HFR tissue using enzymatic tissue disaggregation protocol. **g,** Cardiac NMs from HFR (n=3) proliferate faster compared to those from HF (n=3) tissue. **h,** IF staining of NMs from HF and HFR tissue in culture at day 3, day 7 and day 14 show difference in growth patterns and cell types. **i,** NMs from HFR form vascular like structures by day 14 in culture which stain for EC marker (CD144, Green) surrounded by fibroblasts (Col1a, Red) and dual staining (yellow) cells on edges (arrow). EC-Endothelial Cell, FB-Fibroblast, HF-Heart Failure, HFR-Heart Failure Recovery, IF-Immunofluorescence; LVAD-Left Ventricular Assist Device, LPF-Low Power Field, NM-Non-myocyte (b, c, e-n=5 patient tissue/group, p value-*<0.05, **<0.01, ****<0.0001, using an unpaired t-test; d-Pearson correlation co-efficient test; All graphs show mean ± S.E.M).

### Mechanical Unloading Alters Cellular Proliferation and Plasticity in Cardiac Non-myocytes

Cardiac NMs are primarily ECs, FBs, and mural cells (MCs)^28^. We hypothesize that the myocardial fibrosis in HF is associated with an increase in myocardial FBs and/or MCs with a reduction in EC count. We further hypothesize that, this process is reversed during HFR^19^. To investigate this interplay between ECs and FB/MCs during HFR, we disaggregated fresh human myocardial tissue and isolated cardiac NMs for cell culture in endothelial growth medium (EGM) (Fig. 1f). These studies revealed striking differences between HF and HFR NMs which have not been previously reported. Specifically, in the HF NMs, cell proliferation was impaired, by comparison to the higher cell proliferation rate of HFR NMs, as quantified by electric cell substrate impedance sensing (Fig. 1g). We cultured NMs from both passage 0 and passage 1 and observed similar results. In cell culture, we observed that NMs from HF grew poorly and exhibited a fibroblastic phenotype while cells from HFR reached confluence faster and exhibited an angiogenic phenotype forming vascular network-like structures not observed in HF NMs (Fig. 1h). The vascular-like structures expressed EC markers (CD144/VE-cadherin) and were surrounded by cells expressing FB/MC markers (Col1a2) with dual staining cells on the edges of the structures (Fig. 1i). These studies provided evidence that post-LVAD NMs have greater proliferative capacity, greater plasticity, and a more angiogenic phenotype than pre-LVAD NMs.

### Single-nucleus Transcriptome Reveals Cellular Transitions between HF and HFR

Single-nuclei RNA sequencing (snRNAseq) has been successfully applied to characterize the rich cellular diversity and transcriptional changes associated with specific cell types and states in cardiac tissues and diseases^28–31^. We applied these tools to investigate potential cellular mechanisms of recovery from non-ischemic HF with LVAD support. With single nuclei isolation, library preparation and sequencing of human HF (n=5) and HFR samples (n=5), a total of 6993 nuclei were annotated with cell types after integration and batch correction analysis (Fig. 2a, Supplemental Figs. 2a & 2b). The analysis revealed three distinct subpopulations of FB and five of ECs (Fig. 2b & 2c). A shift in the cellular composition was evident, with a significant increase in an endothelial subtype (EC5) and a concurrent decrease in a fibroblast subtype (FB2) in HFR samples, signaling an active biological transition (Fig. 2d). RNA velocity rank analysis indicated a directional transition with a probability of 0.08 from EC5 to FB2 in HF myocardium, and the reverse transition in HFR with a probability of 0.12 (Fig. 2e, Supplemental Figs. 3a & 3b). Pseudo-time trajectory analysis revealed a dynamic process of cellular transition between subsets of ECs and FBs. Supervised mapping traced the evolutionary pathway from EC to FB phenotype in HF, consistent with an endothelial-to-mesenchyme transition (EndoMT). Such an EndoMT transition would support the development of cardiac fibrosis^32^. By contrast, the pseudotime trajectory analysis in HFR was consistent with an FB transition to EC phenotype (Fig. 2f). Such a cell fate transition could promote restoration of the cardiac microvasculature. This reverse transition was associated with reduced expression of fibrosis-associated genes and increased expression of angiogenesis associated genes during HFR in FB2 and EC5 respectively (Supplemental Figs. 3c & 3d).

**Figure 2.**
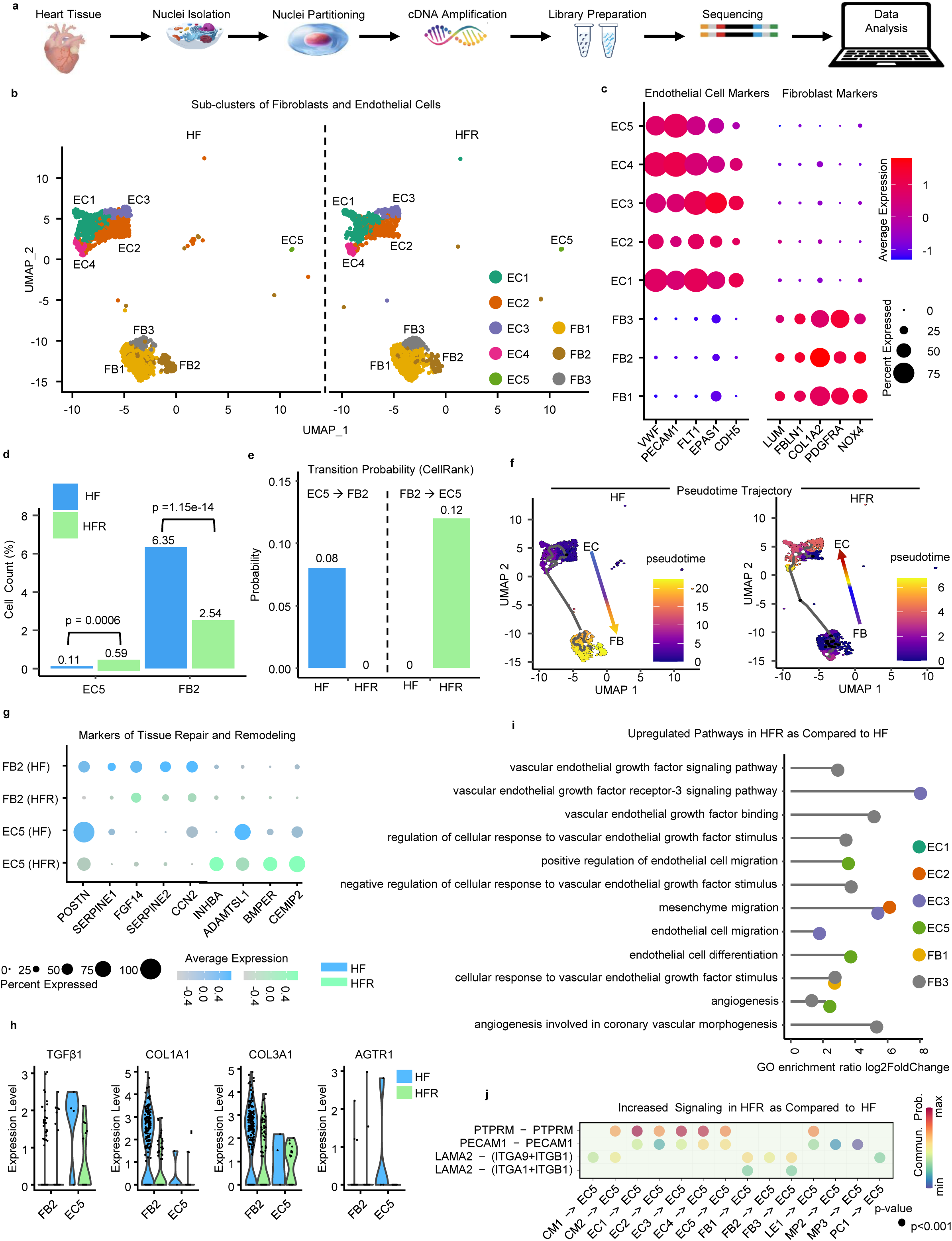
Single-nuclei RNA sequencing suggests mesenchymal to endothelial transition comparing pre-versus post-LVAD cardiac samples. **a,** Schematic overview of the single nuclei RNA-sequencing (snRNA-seq) workflow of HF and HFR cardiac tissue. **b,** UMAP visualization illustrating the heterogeneity of FB and EC populations. **c,** A dot plot showing gene marker expression in subtypes of ECs and FB. **d,** An increase of an EC subtype (EC5) and a decrease of a FB subtype (FB2) in HFR suggesting cellular transition. **e,** RNA velocity analysis indicating dynamic shifts from EC5 to FB2 in HF is reversed in HFR. **f,** A pseudo-time trajectory analysis mapping the transition pathway between FB and EC in HF and HFR samples. **g,** Genes related to tissue repair and remodeling are upregulated in FB2 (HF) and EC5 (HFR), respectively. **h,** The expression of fibrosis-related gene pathways in EC5 and FB2 of HFR is downregulated. **i,** GO enrichment show upregulation in angiogenesis and endothelial cell migration related pathways in ECs and FBs from HFR. **j,** Enhanced ligand-receptor signaling associated with angiogenesis, vascular repair, and ECM remodeling in HFR compared to HF. CM-Cardiomyocytes, EC-Endothelial Cell, ECM-Extracellular Matrix, FB-Fibroblast, GO-Gene Ontology, LE-Lymphatic Endothelial Cell, LVAD-Left Ventricular Assist Device, MP-Macrophages, PC-Pericytes, UMAP-Uniform Manifold Approximation and Projection. (d-two proportion Z-test; j-CellChat default method for the permutation test to calculate significant communication).

Additionally, genes associated with fibrosis including connective tissue growth factor (CCN2)^33^, Periostin (POSTN)^34^ and Serpine-1^35^ are highly expressed in HF in the EC5 subset (Fig. 2g). By contrast, pathway enrichment analysis identified downregulation of fibrotic pathways and diminished expression of fibrosis markers like Transforming Growth Factor Beta 1 (TGFβ1) and the Angiotensin II Type 1 receptor (AGTR1) in the EC5 subset during HFR (Fig. 2h). In addition, genes associated with tissue remodeling and cardiac development such as A Disintegrin and Metalloproteinase with Thrombospondin motif (ADAMTSL1)^36^, BMP-binding endothelial regulator (BMPER)^37^ and cell migration inducing hyaluronidase 2 (CEMIP2)^38^ show increased expression in HFR in the EC5 subset (Fig. 2g). Gene Ontology (GO) enrichment analysis further indicated increased activity of angiogenic pathways and EC migration in HFR (Fig. 2i, Supplemental Fig. 4 & 5). This was complemented by an enhancement in multiple ligand-receptor signaling pathways involved in angiogenesis, vascular repair, and ECM remodeling in HFR (Fig. 2j). For instance, increased FB2 to EC5 ligand-receptor signaling in HFR included laminin subunit alpha 2 (LAMA2) (an ECM protein) and its corresponding ligands, the integrins (ITGA and ITGB), which are involved in cell-ECM adhesion, cell survival and migration.^39^ Altogether, the single-cell transcriptional studies were consistent with an endothelial cell fate transition and reduced fibrosis during HFR.

### Endothelial Cells Transition between Subsets during Recovery

The partition-based graph abstraction (PAGA) velocity analysis and pseudo-time trajectory mapping revealed a shift of EC5 toward other endothelial states EC2, EC3 and EC4 during HFR and a shift from EC2 towards EC1 subset (Fig. 3a-d, Supplemental Fig. 6). To understand the significance of these transitions, we compared gene expressions of the subsets. Notably, endothelial growth factor and angiogenesis-related pathways involved in vascular remodeling and cardiac repair^40^ were upregulated in all the EC subtypes comparative to EC5 (Fig. 3e). These data were consistent with the notion that the EC5 subset may represent a less mature and transitional EC state, whereas other ECs are more mature states (Supplemental Fig. 7-10). In this regard, it is known that the protein tyrosine phosphatase receptor type M (PTPRM) is expressed in endothelial cells of all major organ systems, regulating barrier integrity and mechanotransduction^41^. Furthermore, it is accepted that PECAM1^42^ (CD31), a protein abundant in ECs, plays a key role in inter-endothelial cell junctions and angiogenesis. With respect to these two proteins, we noted an increase in ligand-receptor signaling in the transition from EC5 to other ECs in HFR for both PTPRM and PECAM1 (Fig. 3f). This observation may be consistent with a transition to a more stable EC state (EC1). In this regard, cadherin 5 (CDH5)^43^ and claudin 5 (CLDN5)^44^ (endothelial cell adhesion molecules maintaining a stable endothelial barrier) were more highly expressed in EC1 vs EC5 or EC2 (Fig. 3g). Each of the subsets (EC 1-5) expressed endothelial identity genes such as histone-lysine N-methyltransferase (MECOM) which is crucial for endothelial cell differentiation^45, 46^ (Fig. 3g, Supplemental Fig. 11).

**Figure 3.**
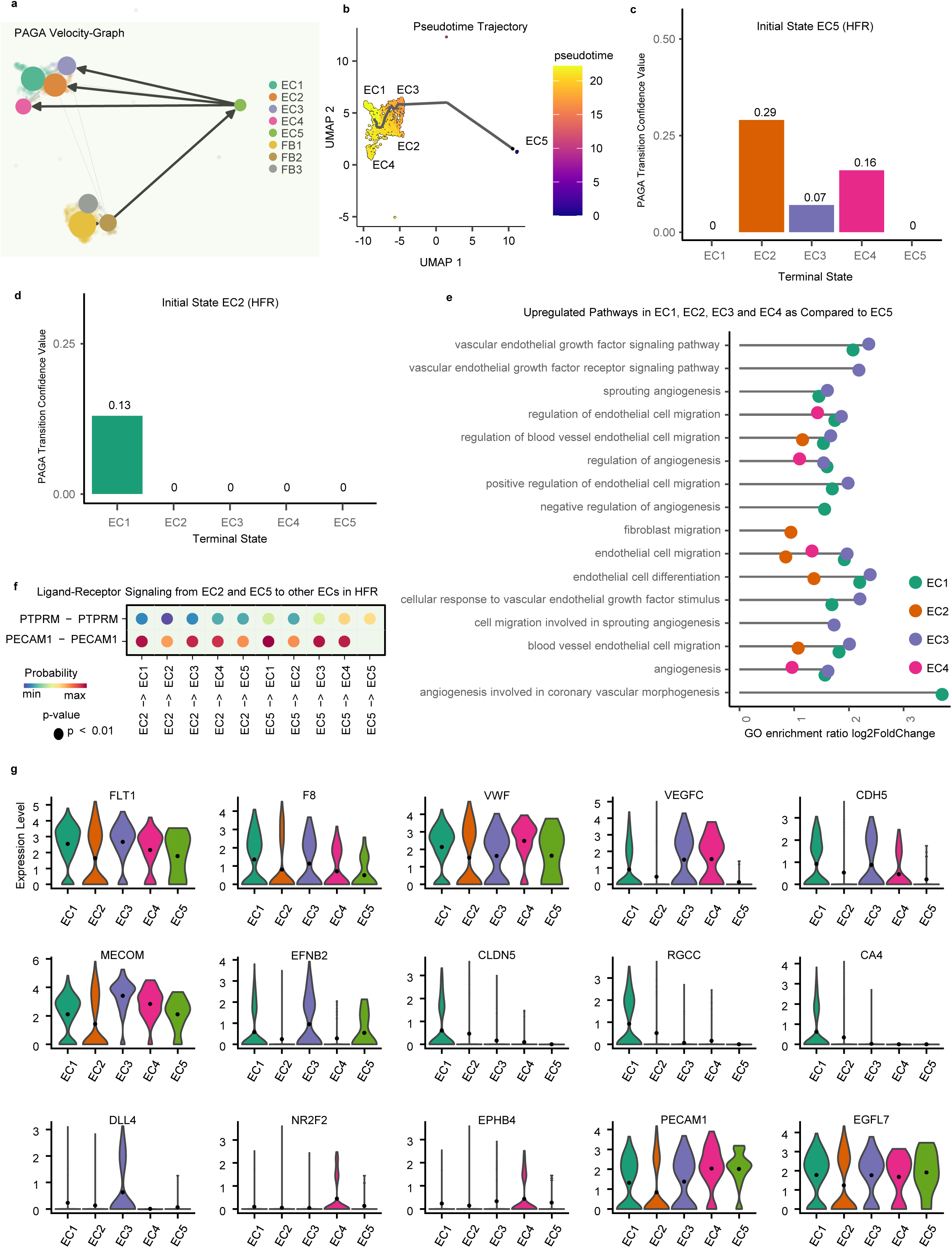
Tracing the fibroblast-endothelial transition via cellular dynamics and gene expression in endothelial cells in HFR. **a**, PAGA velocity analysis suggests a transition probability from the EC5 subtype to other EC subtypes in HFR. **b,** Pseudo-time trajectory outlines a developmental pathway connecting EC5 to other EC subtypes during recovery. **c,** PAGA transition confidence score supports a shift from EC5 towards EC2, EC3 and EC4 in HFR. **d,** PAGA transition confidence score supports a shift from EC2 towards EC1 in HFR. **e,** GO enrichment analysis shows upregulated angiogenesis and endothelial migration-related pathways in all the EC subtypes relative to EC5 in HFR. **f,** Enhanced ligand-receptor signaling associated with angiogenesis and vascular repair in ECs of HFR. **g,** An upregulation is observed in the gene expression of angiogenesis and endothelial lineage related pathways in all EC subtypes (EC1 to EC4) compared to EC5 in HFR. CA4-Carbonic Anhydrase 4, CDH5-Cadherin 5, CLDN5-Claudin 5, DLL4-Delta-Like 4, EC-Endothelial Cell, EFNB2-Ephrin B2, EGFL7-Epidermal Growth Factor-Like Protein 7, EPHB4-Ephrin Type-B Receptor 4, F8-Coagulation Factor VIII, FLT1-Fms related receptor tyrosinase kinase 1, GO-Gene Ontology, LVAD-Left Ventricular Assist Device, MECOM-MDS1 and EVI1 Complex Locus, NR2F2-Nuclear Receptor Subfamily 2 Group F Member 2, PAGA-Partition based Graph Abstraction, PECAM1-Platelet and Endothelial Cell Adhesion Molecule, RGCC-Regulator of Cell Cycle, VEGFC-Vascular Endothelial Growth Factor C, VWF-Von Willebrand Factor. (f-CellChat default method for the permutation test to calculate significant communication)

### Murine Model of Heart Failure and Recovery

Our group has established a mouse model of non-ischemic HF and recovery^19^. Briefly, HF is induced by oral administration of high salt and L-N^G^-Nitro arginine methyl ester (L-NAME) together with subcutaneous administration of angiotensin II via an osmotic pump over 5 weeks (Fig. 4a). After cessation of this HF induction protocol, heart failure recovery (HFR) occurs over the ensuing 4 weeks as assessed by echocardiography and histology.

**Figure 4.**
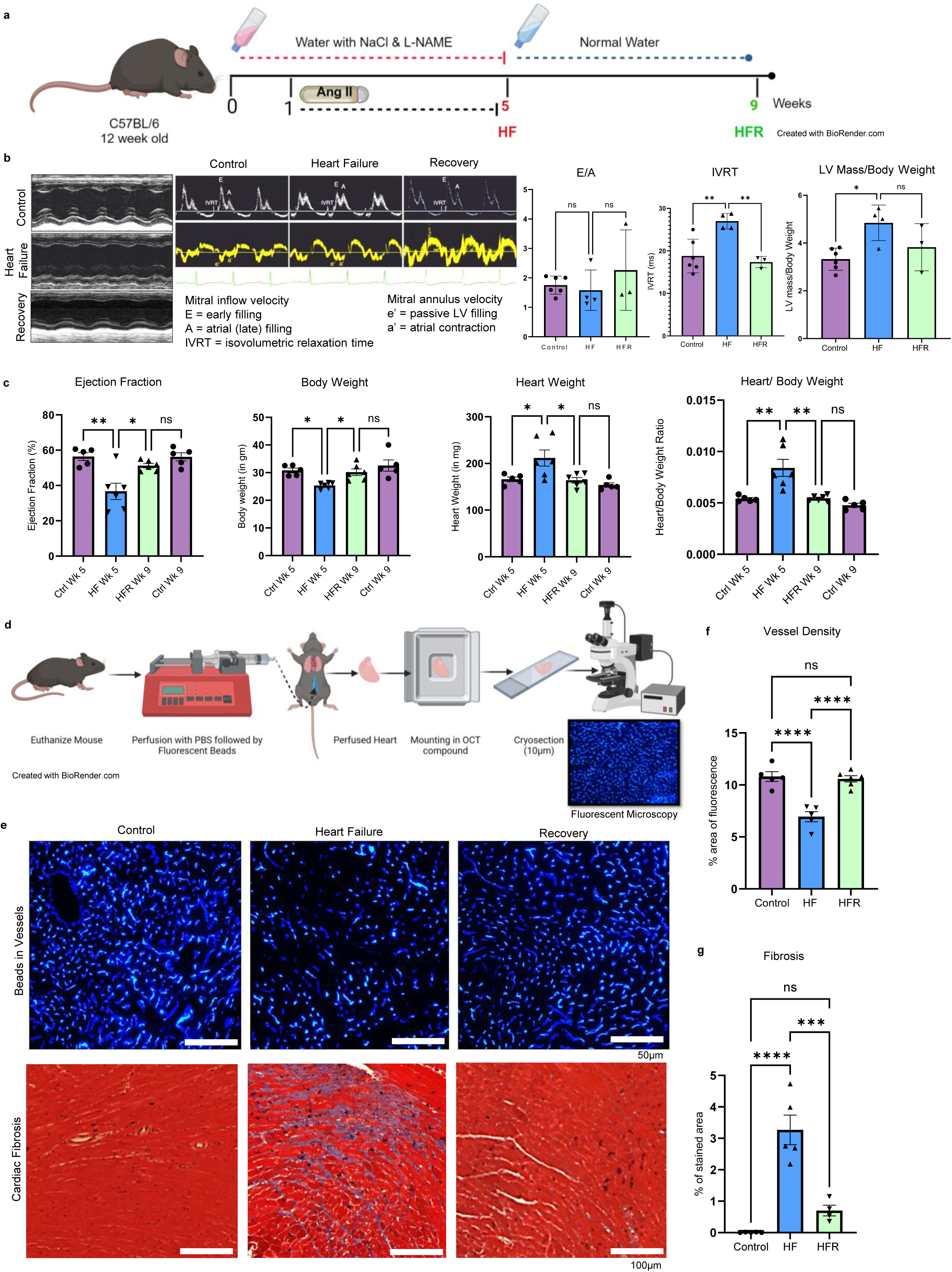
Non-ischemic mouse model of heart failure and recovery recapitulates human heart failure and recovery. **a**, Schematic of non-ischemic mouse model of heart failure and recovery. **b,** Representative echocardiography images of ejection fraction, E/A ratio, isovolumetric relaxation time in heart failure and recovery mice; HFR mice show improved E/A, reduced IVRT and LV mass/body weight. **c,** Recovery from heart failure based on EF, heart weight and body weight ratio. **d**, Schematic of fluorescent microsphere bead perfusion technique to assess vascular density. **e,** Blue fluorescent microsphere beads in vessels to assess vascular density and Masson’s trichrome staining to assess fibrosis. **f&g,** Vessel density increased along with reduction in fibrosis in HFR mice. EF-Ejection Fraction, HFR-Heart Failure Recovery, IVRT-Isovolumetric Relaxation Time, LV-Left Ventricle. (b, Control-n=6, HF-n=4, HFR-n=3; c, Age matched controls-n=5, HF-n=6, HFR-n=6; f, Control-n=5, HF-n=5, HFR-n=6; g, Control-n=5, HF-n=5, HFR-n=4 p value-*<0.05, **<0.01, ***<0.005, ****<0.0001, using one-way ANOVA, All graphs show mean ± S.E.M)

In the current study, the pharmacological induction of HF was manifested at week 5 by a reduction in ejection fraction (from 56 ± 4.8% to 36 ± 11%, *p*=0.001), and in the E/A ratio (from 1.8 ± 0.3 to 1.6 ± 0.7); together with an increase in isovolumetric relaxation time (IVRT; from 18.8 ± 3.9 to 26.9 ± 1.8ms, *p*=0.004), left ventricular mass to body weight ratio (from 3.3 ± 0.5 to 4.8 ± 0.7, *p*=0.01); and heart weight (from 166 ± 11 to 212 ± 42 mg, *p*=0.03; Figs 4b and 4c). After cessation of the drugs in week 5, these physiological and echocardiographic features of HF normalized by week 9 (HFR; Figs. 4b & 4c, Supplemental video).

### Recovery from Heart Failure is a Vascular Recovery, in part due to the MEndoT

To investigate whether HFR is associated with vascular recovery, we quantified vascular density in mice using fluorescent microsphere bead perfusion (Fig. 4d). In HF mice, there was a reduction in percent area of fluorescence (6.9 ± 1%) compared to control mice (10.8 ± 1%), *p*<0.0001. By contrast HFR mice manifested a restoration of fluorescence area (10.5 ± 0.7%), *p*<0.0001 (Fig. 4e & 4f, Supplemental Fig. 12). These studies indicated that vascular density was reduced in HF but restored in HFR. Fibrosis was quantified using Masson’s trichrome staining. Fibrosis increased in HF (control vs HF p<0.0001) whereas during HFR, fibrosis decreased (HF vs HFR *p*=0.0003) (Fig. 4e and 4g). Cardiac tissue fibrosis in HF and HFR mice mirrors changes seen in human pre-LVAD and post-LVAD cardiac tissue samples. The increase in vessel density in HFR mice suggests a vascular component of recovery.

To evaluate angiogenic transdifferentiation as a potential mechanism for HFR, we used tamoxifen inducible fibroblast fate-mapped Col1a2-creERT2: R26R^tdTomato^ mice. Col1a2 has been reported as a reliable marker to identify cardiac FBs in the adult mouse heart^47–49^. To induce Col1a2-cre tdTomato labeling, tamoxifen was administered at the end of HF induction (5-week timepoint) for five consecutive days (Fig. 5a). Subsequently, hearts were harvested at 9 weeks (at a time of recovery from heart failure, i.e., HFR) for immunohistochemistry and flow cytometry analysis (Fig. 5a, Supplemental Fig.13). In the animals subjected to the HF protocol and allowed to recover, a proportion of the tdTomato+ cells co-expressed the endothelial marker CD31/PECAM-1 by immunohistochemistry (Fig. 5b & 5c). These findings are consistent with transdifferentiation of some fibroblasts to endothelial cells. These observations were confirmed and quantified by flow cytometry, where the proportion of transdifferentiating cells was 5.6% in the HFR group vs. 0.4% in the control group, *p*<0.0001 (Fig. 5d & 5e, Supplemental Fig. 14). Here it is important to note that our fibroblast labeling strategy probably did not identify all cardiac fibroblasts as only 17% of NMs in control mice and 24% in HFR mice were tdTomato+ (fibroblasts should make up at least 35% of NMs) (Supplemental Fig. 14). Accordingly, the percentage of transdifferentiating cells is likely an underestimate. In any event, the evidence supports an angiogenic transdifferentiation of cardiac fibroblasts takes place during recovery from HF.

**Figure 5.**
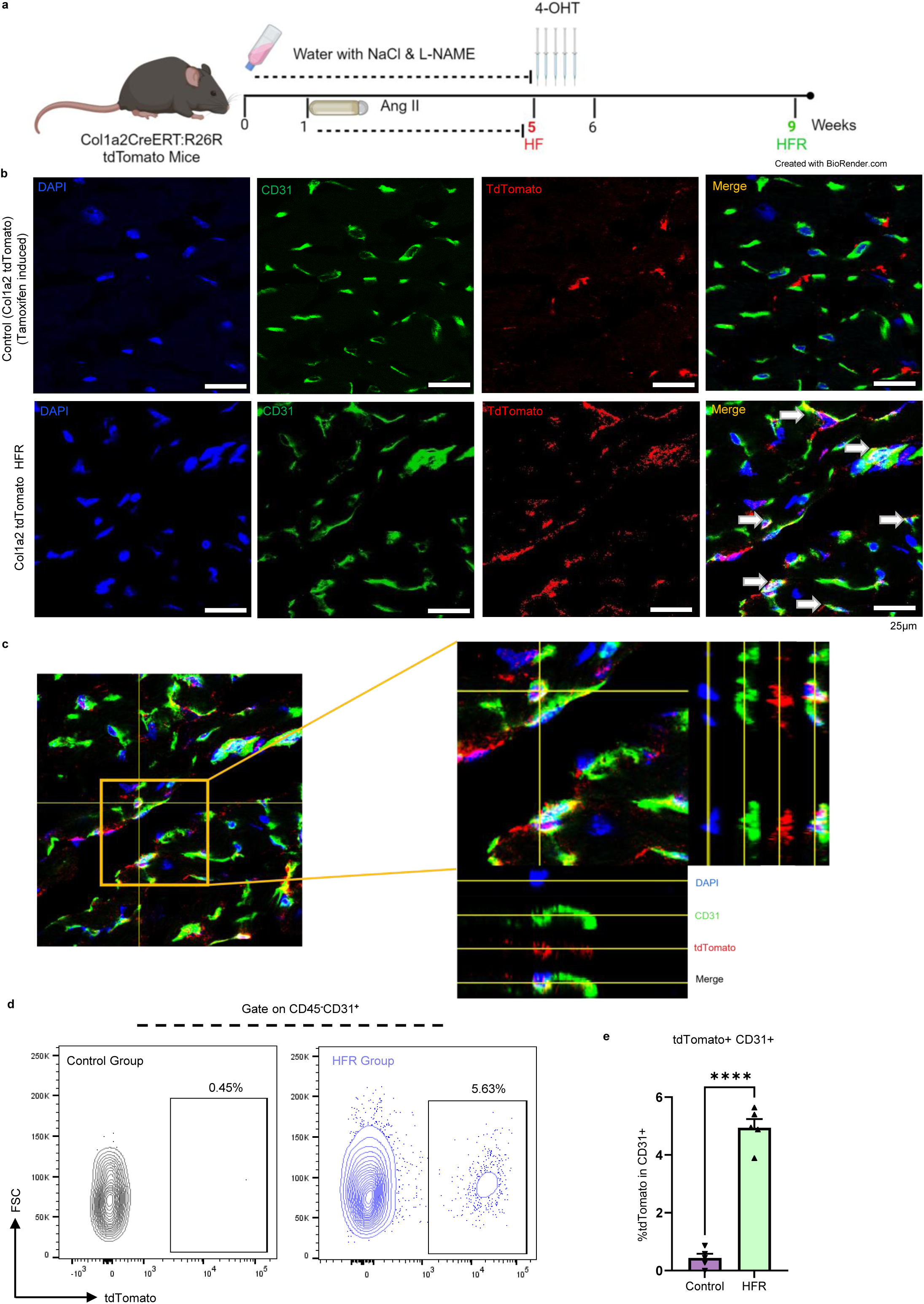
Angiogenic transdifferentiation during heart failure recovery in Col1a2-CreERT:R26R tdTomato mice. **a**, Schematic of fibroblast lineage tracing in the HFR mouse model. Col1a2-expressing fibroblast cells and their progenies are marked with tdTomato after tamoxifen (4-OHT) induction. **b,** IF staining visualized by confocal imaging in HFR group showed CD31+ (green) tdTomato+ (red) dual staining (yellow) cells representing endothelial cells transdifferentiated from fibroblasts (arrows - dual staining cells). **c,** The 3-side view by confocal microscope confirmed the dual staining of the cells that expressed both CD31 and tdTomato. **d,** Angiogenic transdifferentiation during HFR quantified by flow cytometry. **e,** HFR group shows increased number of transdifferentiated (CD31+ tdTomato+) cells. Control-Non-operated tamoxifen induced age-matched mice; HFR-Heart failure recovery. (n=5 mice/group, p value-****<0.0001, using an unpaired t-test, Graph shows mean ± S.E.M)

## Discussion

The current study indicates that the recovery from heart failure (HF) is associated with an expansion of the microvasculature to improve cardiac perfusion, mediated in part by angiogenic transdifferentiation. We compared histological, molecular, and cellular attributes of human cardiac samples obtained at the time of LVAD implantation (pre-LVAD or HF) or at LVAD explantation (post-LVAD or HFR). Specifically, we find that non-myocytes isolated from HF patients manifest an impairment in cell proliferation and plasticity, with a fibroblastic phenotype. By contrast, cardiac NMs derived from patients on chronic LVAD support have improved cell proliferation and plasticity, spontaneously forming vascular networks in cell culture. Furthermore, in pre-LVAD specimens, we observed a transcriptional signature and RNA velocity consistent with an endothelial-to-mesenchyme transition, whereas in post-LVAD hearts, the transcriptional signature and RNA velocity were consistent with a MEndoT. In a murine model of HF, we observed interstitial fibrosis, reduced capillary density and vascular volume, with impaired cardiac function and structure. By contrast, in HFR we observed a reduction in interstitial fibrosis and FB number, with an increase in capillary density and vascular volume, and an improvement in cardiac structure and function. Lineage tracing studies revealed that the increase in capillary density was associated with a fibroblast-to-endothelial cell fate transition that we have termed angiogenic transdifferentiation^50^.

The approach to improving cardiac function in left ventricular failure has focused on afterload reduction and heart rate control (to reduce the work of the heart); pre-load reduction (to reduce pulmonary congestion); control of arrhythmias; inhibition of adverse (and augmentation of favorable) neurohormonal influences (i.e., antagonists of the sympathetic nervous system, the renin-angiotensin axis, and neutral endopeptidase); and metabolic control as with SGLT2 inhibitors. Positive inotropic support is useful during acute decompensation, but long-term inotropic activation has not been useful^51^. Methods to increase myocyte proliferation or to enhance myocardial metabolism are experimental^52, 53^.

Although strategies of HF treatment have largely focused on myocytes, it is increasingly apparent that NMs contribute to HF. The disseminated interstitial fibrosis that is ubiquitous in “non-ischemic” HF was long thought to be irremediable, but recent observations suggest otherwise^54–56^. In particular, HF patients receiving LVAD often manifest some improvement in cardiac function, associated with a reduction in cardiac fibrosis and an increase in capillary density. This is particularly true in LVAD patients with non-ischemic HF^57, 58^. There may be an important role for a vascular niche and non-cardiomyocyte signaling in recovery from HF^40, 59^. Indeed, our study reveals a novel interaction between FBs and ECs of the recovering heart, with a downregulation of pro-fibrotic and an upregulation of pro-angiogenic genes. Studying the mechanism of cardiac recovery on LVAD support can provide valuable insights into the mechanisms underlying endogenous cardiac tissue regeneration, repair, and recovery.

Our detailed snRNA-seq analyses have revealed an important interplay between cellular transitions and gene expression alterations within the cardiac tissue microenvironment. Notably, we observed a downregulation of fibrosis markers/mediators, including COL1A1, COL3A1, TGF-β1 and AGTR1^60–62^ particularly in the EC5 subset. As each of these genes play a role in the accumulation of ECM, and their reduced expression in the post-LVAD specimens may facilitate a recovery from cardiac fibrosis. Interestingly, we found the upregulated expression of inhibin beta A (INHBA) in EC5 subset of HFR. Traditionally, INHBA^63^ has been associated with epithelial-mesenchymal transitions but recently Dogra *et al*.^64^, reported a role for INHBA upregulation in cardiac regeneration which is consistent with our work. Furthermore, we observed an upregulation of angiogenic genes following LVAD implantation, particularly in the EC5 subset. This dynamic shift suggests a transition away from fibrotic pathology toward an angiogenic and reparative cellular milieu.

In addition, the PAGA velocity analysis revealed a transition from one EC subset (EC5) to other EC subtypes as well as a transition from EC2 to EC1 in HFR, the latter subset having a more mature EC transcriptional profile. For example, EC1 subset manifested greater expression of PECAM1 which plays a key role in inter-endothelial cell junctions and angiogenesis^42^. Similarly, the increased expression of PTPRM in EC1 is notable in that this gene promotes cell fate transition in cervical cancer^65^, and could have a similar role in cardiac cell fate transitions. The increased expression of endothelial markers-such as vWF^66^, FLT1^67^, and EGFL7^68^ in all EC subtypes compared to EC5 in HFR is also concordant with a shift toward a more mature EC state during angiogenic transdifferentiation^69–71^.

Our cell culture experiments, utilizing freshly isolated cardiac NMs from HF and recovery tissues, provide compelling evidence of transitioning cell populations. Specifically, we have discovered a greater proliferative capacity of NMs during recovery and their potential to form vascular networks. These findings are consistent with the sn-RNA seq data. While snRNA-seq studies offer a snapshot in time, cell culture experiments permit observation of dynamic changes over time. In the proliferating and evolving cell population in culture, we can explore cell fate and unravel complex cell-to-cell interactions. Future investigations will delve deeper into the mechanisms underlying these cellular adaptations, and may be a model for guiding the development of novel therapeutics for HF.

We employed a non-ischemic mouse model of HF and recovery to confirm the vascular nature of HFR, and to explore mechanisms. This model closely recapitulates the observations from human HF and HFR tissues, including the changes in cardiac function and histology. We employed microsphere bead perfusion techniques to confirm an increase in vascular volume during HFR.

To further elucidate the origin of the transdifferentiating cell population, we employed the Col1a2-creERT2: R26R^tdTomato^ mouse model. The dual staining FBs during HFR expressed an endothelial marker CD31 and tdTomato, confirmed the MEndoT. Of note, only 5.6% of cells exhibited angiogenic transdifferentiation at the time point measured. This is likely an underestimate of the cardiac FBs undergoing transition to ECs. Specifically, our FB labeling efficiency was incomplete as FBs should make up at least 35% of NMs (whereas only 17% in control mice and 23.6% in recovery mice, of NMs were labeled by tdTomato). Accordingly, the fibroblast-derived ECs (tdTomato+CD31+) identified by our labeling strategy are likely an under-representation of the true prevalence of angiogenic transdifferentiation.

He *et al*.^72^, did not find evidence of MEndoT in their meticulous lineage tracing studies in a murine model of myocardial ischemia and reperfusion. They observed that cardiac FBs expand substantially after the injury, but do not contribute to the formation of new coronary blood vessels, indicating no contribution of MEndoT to neovascularization. This model of myocardial injury is quite different from our murine model of *non-ischemic* cardiomyopathy. In this regard, it is intriguing to speculate that, in patients with HF due to *ischemic* cardiomyopathy, there may be a lack of MEndoT. This hypothesis, based on the murine model of He *et al*., might explain the observation that HF patients with *ischemic* cardiomyopathy are much less likely to recover post-LVAD implantation.

By contrast, Dong *et al.*^73^, observed MEndoT during the onset of cardiac hypertrophy in a murine model of transaortic constriction. It is possible that in this murine model, MEndoT and increased neovascularization is a compensatory response to the increased cardiac afterload of transaortic constriction. This model is quite different from that of our murine model of HFR, which mimics the histological and functional changes that occur in the human heart post-LVAD.

In conclusion, we propose that recovery from heart failure is a **microvascular recovery**. This microvascular recovery is in part due to an angiogenic transdifferentiation of a subset of fibroblasts to endothelial cells. This new paradigm may lead to a novel therapeutic avenue for heart failure treatment.

## Methods

### Human Myocardial Samples and Experiments

#### Tissue Acquisition

Cardiac tissue samples were collected at the time of the LVAD implant (n=8) or after explant (n=8). The core of tissue removed at the time of LVAD implantation from the apical region of the heart was used (pre-LVAD); and the left ventricle free wall adjacent to the apical region was used for post-LVAD samples at the time of cardiac transplantation. In both cases, only midmyocardial region of the tissue was sampled (endocardium and epicardium excised and discarded) (Figure 1A). Cardiac tissue samples were acquired based on an institutional protocol approved by the Houston Methodist Hospital Institutional Review Board [IRB Pro 00006097] and abide by all ethical regulations corresponding to human tissue research. Tissues were collected and immediately processed for freezing and paraffin embedding for histology. Tissues were flash-frozen at -80℃ for protein and RNA assays. For histology, tissues were promptly fixed in 4% paraformaldehyde. Subsequently, the samples were dehydrated in a series of graded alcohols, cleared in xylene, and embedded in paraffin following standard techniques. 5µm thick sections were cut for histology. For snRNA sequencing analysis, frozen samples from five randomly selected patients from the HF and HFR groups were each pooled and processed as HF and HFR human tissue grouped sample.

#### Cardiac Fibrosis

To quantify fibrosis, sections were stained using Masson’s trichrome (Sigma-Aldrich, HT15-1KT), following the manufacturer’s instructions. Slides were cover-slipped and examined under 10x objective using an Olympus BX61 microscope. Whole tissue sections of the heart were photographed, and fibrosis was quantified using Olympus cellSens dimension software. A user-independent computerized color cube-based selection criterion, focusing on the color spectrum of the blue dye, was applied to denote positive staining. Stained and unstained areas were measured, expressing the results as the average percentage of tissue (pixels) stained by the dye. The analysis was conducted by a single investigator who was blinded to the samples.

#### Fibroblast and Endothelial Cell Count

Myocardial fibroblasts and endothelial cells were quantified by staining for antibodies vimentin AF647 (1:200, sc-32322, Santa Cruz) and CD144 AF647 (1:200, sc-9989, Santa Cruz), respectively. Five microscope fields (20x) were analyzed for quantification and the results were expressed as mean ± SD. The analysis was done by a single investigator blinded to the source of the samples. Additionally, von Willebrand Factor rabbit polyclonal (1: 200, ab9378, Abcam) with donkey anti-rabbit 555nm secondary ab (1:500, A31572, Invitrogen) was used to stain ECs and capillaries.

#### Cardiac Non-myocyte Cell Isolation

Freshly received cardiac tissue underwent sequential enzymatic tissue digestion using DMEM+Collagenase (10:1) (Liberase TM Research Grade, 5401127001, Roche Diagnostics) at 37℃. After disaggregation, the solution was filtered and differentially centrifuged to obtain a pellet of cardiac NMs. The resulting cell pellet was resuspended in EGM2 (Endothelial Cell Growth Medium-2 Bulletkit^TM^, CC-3162, Lonza, Walkersville, MD USA) and divided for freezing and cell culture. For future assays, cells were frozen at -80℃ in equal amounts of freezing media (Cell Freezing Medium-DMSO 1x, Sigma-Aldrich, C6164). The other half was plated for cell culture and immunofluorescence assay on coverslips placed in 6-well culture plates. Freshly isolated cells are referred to as P0 (passage 0) and the first passage of cells are labeled as P1 once confluent.

#### Electric Cell-substrate Impedance Sensing Assay (ECIS)

The isolated cells, frozen at P1 were thawed from HF and HFR samples (n=3). The cells were then plated in duplicates on two ECIS 8-well chambered slides (8W10E+PET, Applied Biophysics, Inc.) and cultured in EGM2. The plates were attached to the ECIS machine (The ECIS Z-Theta, Applied Biophysics, Inc. Troy, NY 12180) and their resistance/ohm was measured until the cells reached a confluent monolayer indicated by plateau on the graph. EGM2 was changed after the first 24 hours and subsequently every 3 days.

#### Immunofluorescence

The coverslips from culture plates were stained on Days 3, 7 and 14. Coverslips were immune-stained with conjugated primary antibodies for endothelial cells - CD144 AF647 (1:200, sc-9989, Santa Cruz) and for fibroblasts - Col1a AF647 (1:200, 1310-31, Southern Biotech). The stained coverslips were then inverted and mounted on slides and imaged with a fluorescence microscope (Olympus BX61 Fluorescence Microscope) at 10x, 20x and 40x objective lens. Z-stacks were also taken using a confocal microscope (Olympus FV3000).

#### Single Nuclei Isolation and Library Preparation for Sn-RNA Sequencing

We isolated nuclei from frozen human heart tissue samples and constructed 3’ single-cell gene expression libraries (Next GEM v3.1) using the 10x Genomics chromium system. The libraries were sequenced with ∼200 million PE150 reads per sample on Illumina NovaSeq platform. After sequencing, we mapped the raw reads of RNA sequencing data to the human reference genome (GRCh38) using Cell Ranger (version 7.1.0) and counted the unique molecular identifiers (UMIs) for each gene. We then loaded the resulting UMI count matrix into the Seurat R package (version 4.3.0)^74^. We retained high-quality cells that expressed between 200 and 2500 genes, excluding those with more than 5% mitochondrial gene content. Genes expressed in fewer than three cells were also filtered out. After filtering, the high-quality data were normalized, with cell-level scaling to 10,000 reads and log-transformed after incrementing counts by one. The normalized data underwent further processing steps: scaling (scale data function), principal component analysis (PCA) (RunPCA function, npcs =30), Uniform Manifold Approximation and Projection (UMAP) (Run UMAP function, reduction = “pca”, dims = 1:30), shared nearest neighbor graph (SNNG) construction (Find Neighbors function, reduction = “pca”, dims = 1:30), and cell clustering (Find Cluster function, resolution=0.8).

##### Sn-RNAseq Differential Expression and Cell Type Annotation

Furthermore, we conducted Differentially Expressed Genes (DEGs) analysis in one cluster versus other clusters using the Find All Markers function. The Wilcoxon test method was used by default, with a minimum percentage of expressed cells set to 25% and a minimum log2 fold change of 0.25. Cell types were annotated based on known marker genes from PanglaoDB^75^, cell-Taxonomy^76^, disco^77^ databases, and relevant literature. We kept FBs, ECs, and their subpopulations for further analysis, except for cell-cell communication. Marker gene expressions were visualized by Dot Plot and Vln Plot functions.

##### Cell Fate Transition, Trajectories, and Cell-Cell Communication with Functional Enrichment Analysis

For cell fate transition, trajectory analysis, and cell rank for directed single-cell transition mapping, we utilized scanpy (version 1.10.2)^78^, scVelo (version 0.2.5)^79–81^, Monocle3 (version 1.3.7)^82–85^ and CellRank (version 1.5.1)^86^ with default parameters. For trajectory analysis, FBs in post-LVAD and ECs in pre-LVAD were set as initial cell types. We used the cellrank.tl.terminal _state and cellrank.tl.initial_state functions with n_states=5 to estimate the initial and terminal state of directed single-cell transition mapping. Transition probability and changes in cell proportion analysis between the conditions were visualized by bar plots. Changes in cell proportions between conditions were tested for statistical significance employing the function (prop.test). The ligand-receptor cell-cell communication was analyzed by CellChat (version 1.5.0)^87^. The communication probability of cell-cell communication was analyzed using circle plots. Finally, Gene Ontology (GO) functional enrichment analysis was performed using the clusterProfiler R package (version 3.18.1)^88^ and visualized by bar, point, tile and cnet plots.

#### Animals and Induction of Heart Failure

All animal experiments were conducted with approval from the Houston Methodist Research Institute Institutional Animal Care and Use Committee (Houston, TX) and were in accordance with the guidelines for the Care and Use of Laboratory Animals published by the US National Institutes of Health (NIH Publication No. 85-23, revised 1985). 2-month-old male wildtype C57BL/6 mice were bought from Envigo, Alice, TX and were individually housed under 12-h:12-h light/dark cycle with enrichment and acclimatized for 3 days. Col1a2-creERT2 and R26RtdTomato were purchased from The Jackson Laboratory (Bar Harbor, ME) to generate Col1a2-creERT2: R26R^tdTomato^ mice, Col1a2-creERT2 mice were crossed with R26R^tdTomato^ mice^89^. Mice were planned n=5 per group based on extensive previous work with this model for statistical significance. Mice which reached a pre-identified euthanasia point were removed from the experimental study. Experimental groups were identified using sequential numerical cage numbers as placed by husbandry staff.

#### A Mouse Model of Heart Failure and Recovery

Details on the murine HF model previously established by our group are described earlier^19^. In brief, HF was induced in 3-month-old C57BL/6 or Col1a2-creERT2: R26R^tdTomato^ mice by adding 0.3mg/mL L-NAME (Sigma-Aldrich, St. Louis, USA) and 1% NaCl (Sigma-Aldrich, St. Louis, USA) to the drinking water for 5 weeks. One week after starting water, angiotensin-II (Sigma-Aldrich, St. Louis, USA) was delivered through subcutaneous osmotic pumps (DURECT Corporation, Cupertino, USA) at a rate of 0.7 mg/kg per day for 4 weeks (Fig. 4a). To induce Col1a2-cre tdTomato labeling, 1mg of Z)-4-hydroxytamoxifen (Sigma-Aldrich, St. Louis, USA) was administered by intra-peritoneal injection at the end of HF induction (5-week timepoint) for five consecutive days (Fig. 5a). Subsequently, hearts were harvested at 9 weeks (at the time of recovery from HF, i.e., HFR) for immunohistochemistry and flow cytometry analysis.

#### Echocardiography

Transthoracic echocardiography was performed as previously described^90^. In brief, mice were anesthetized with Isoflurane 1.5 % (AbbVie, North Chicago, IL) and 98.5% O_2_ and placed on a warming pad at 37.5°C. Measurements were performed with the Vevo 1100 imaging system and an MS400 (18-38MHz) transducer. Images were analyzed using Visual Sonics Software Vevo Lab 1.7.1 (Visual Sonics, Toronto, Canada) in a blinded fashion. Systolic function was assessed in the parasternal longitudinal axis (PSLAX) in B-Mode, and ventricular wall dimensions were assessed on papillary muscle level in M-Mode from the parasternal short-axis view. The E/A ratio was assessed in an apical 4-chamber view.

#### Mouse Microsphere Bead Perfusion Study

A modified mouse perfusion method, as described by Springer *et al*.^91^ was employed to perfuse mice with Fluospheres carboxylate-modified 0.2 µm blue microsphere beads (Invitrogen, Catalog no. F8805) diluted at 1:20 with phosphate buffered saline (PBS 1X). Briefly, mice were administered deep inhalation anesthesia, the right ventricle was identified, and a small incision was made to allow excess perfusate to exit the vascular space. Using a 26-gauge needle, the left ventricle was accessed via the apex. The mouse was initially perfused with 8ml of PBS at a rate of 10ml/min using a syringe pump. Subsequently, fluorescent microsphere beads (5ml) were used for perfusion. Upon completion of the perfusion, the heart was immediately removed from the chest cavity, weighed, and placed in OCT (optimal cutting temperature) media for freezing at -80℃. Deep cryosections were cut at 10µm thickness using a cryostat for histology and imaging of the beads’ fluorescence (Fig. 4d).

#### Fluorescent Beads Analysis

Cryosections were imaged under 20X objective using an Olympus BX61 Fluorescence microscope. The DAPI channel was utilized to visualize blue-colored fluorescent beads. Cross-sectional images of the entire heart were captured at the papillary muscle level. A blinded observer analyzed the images for vascular volume representing vessel density using CellSens Dimension Software from Olympus. On each image, a region of interest (ROI) was marked to exclude tissue folds and areas lacking perfusion. The results are expressed as the average percent area (pixels) comprising fluorescent beads. Additionally, CD31 mouse (1:200, NBP2-34283, Novus Biologicals) and AF647 donkey anti-mouse (1:500, A-31571, Invitrogen) staining was performed on some tissue sections to confirm that beads filled the small capillaries without leaking out of the vessels. (Supplemental Fig.12).

#### Cell Isolation and Flow Cytometry

Single cells from freshly isolated mice hearts were isolated according to the manufacturer’s protocol (Miltenyi Biotec, Bergisch Gladbach, Germany). Samples were then Fc blocked with CD16/32 (14-0161-82, Invitrogen) and stained with the following antibodies: CD31-APC (1:200,17-0311-82, Invitrogen), PDGFRa-FITC (1:200, 11-1401-82, Invitrogen), and CD45-PE-Cy7 (1:1000, 25-0451-82, Invitrogen). DAPI (1:400, 564907, BD Biosciences) was used to exclude dead cells. Analysis was performed with assistance from the Houston Methodist Research Institute Flow Cytometry Core on a BD FACS Fortessa machine. Data were analyzed with FCS Express 7 and FlowJo v10.

#### Immunofluorescence Tissue Preparation and Confocal Microscopy

Freshly isolated mice hearts were fixed with 4% PFA for one hour at 4°C and subsequently transferred to 30% sucrose overnight at 4°C and then embedded in OCT. Samples were sectioned at 10µM and stained using the following primary antibodies: anti-mouse CD31 rat (1:50, 550274, BD Pharmingen), RFP Chicken (1:1000, 600-901-379S, Rockland) and AF488 donkey anti-rat (1:200, a21208, Invitrogen) and AF647 goat anti-chicken (1:200, a21449, Invitrogen) as secondary antibodies respectively. The z-stack images were captured using an Olympus confocal microscope (FV3000) with a 40X objective lens. The obtained images were analyzed using cellSens dimension software. Merged signals and split signals were used to delineate the signals for single-cell resolution.

#### Statistical Analysis

The data are presented as mean ± standard deviation (SD). Differences between groups were identified using the Student’s *t*-test (two-tailed) and between multiple groups using one-way or two-way analysis of variance (ANOVA). The Pearson correlation coefficient test was performed to measure linear correlation. A two-tailed *p* <0.05 was used as the significance cut-off for all tests. Analyses were conducted using Graph Pad Prism 9.3.0 software (Graph Pad Software Inc., La Jolla, CA). Graphically, all results are shown as mean ± standard error (SEM).

## Data Availability

All the data supporting the findings in this study are included in the main article and associated files. Source data are provided with this paper. Raw sequencing files and processed normalized data have been submitted to the NCBI Gene Expression Omnibus (GSE253535).

https://www.ncbi.nlm.nih.gov/geo/query/acc.cgi?acc=GSE253535

## Acknowledgements

We thank Houston Methodist Research Institute Flow Cytometry core and Advanced Cellular & Tissue Microscopy core for their assistance in analysis and confocal imaging, respectively. Human tissue was collected under an approved IRB protocol Pro00006097.

## Funding Source

This work was supported by a grant to JPC and KC from the National Institute of Health (RO1 HL148338).

## Author Contributions

RKR performed most of the experiments on human tissue and mice perfusion studies with the assistance of KAY, A.M. contributed with the cell proliferation assay, K.G. analyzed data for RNA -seq with assistance from K.C. and L.Z., L.L. assisted with human RNA-seq, AJL & M.G. performed lineage tracing mouse studies, S.L., F.N. and KNC conducted the analysis, JPC., KAY., A.B., K.C. designed the study, R.R., K.G. wrote drafts of the manuscript and F.N. edited it. All authors contributed to the revisions of the manuscript.

## Conflict of Interest

Authors declare no conflict of interest.

## Code Availability

The token ‘mrmhkguqvrwzjwx’ allows anonymous, read-only access to GSE253535. The code used for the bioinformatic analysis in this study is not publicly available but can be accessed with permission from the corresponding author.

---

**Supplemental Table 1:** Demographics of Pre- and Post-LVAD Patients.

| Demographics | Pre-LVAD (n=8) | Post-LVAD (n=8) |
| --- | --- | --- |
| Age | 55±13 (years) | 51±15 (years) |
| Gender | 50% Male | 75% Male |
| Non-ischemic<br>Cardiomyopathy | 88% | 88% |
| Diabetes Mellitus | 62% | 50% |
| Hypertension | 62% | 75% |
| Hyperlipidemia | 50% | 50% |
| Ejection Fraction | 19.6±3.4 (%) | 28.4±18 (%) |

**Supplemental Figure 1.**
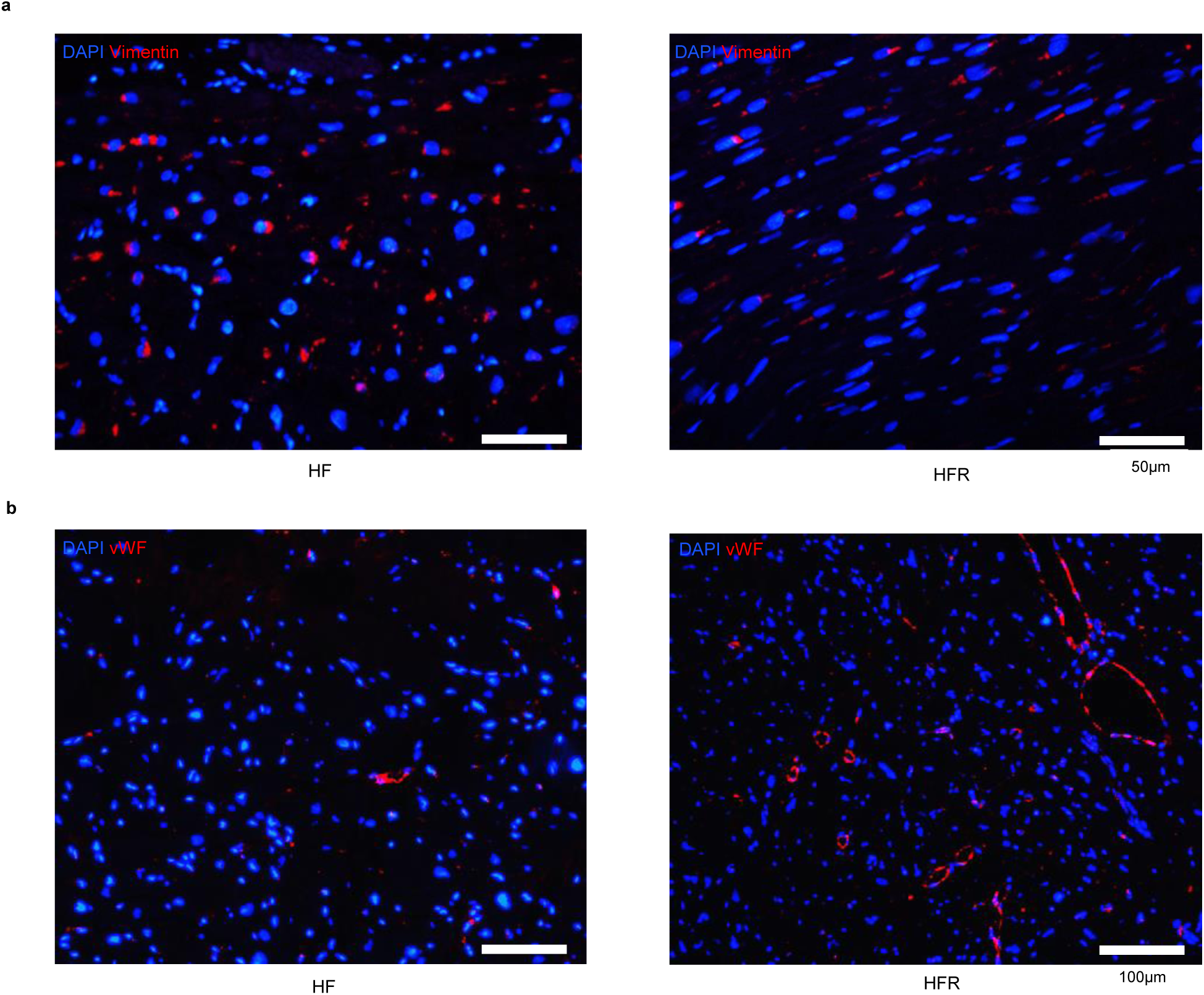
Reduction in fibroblasts and an increase in capillaries in HFR. **a,** Vimentin staining shows decrease in fibroblast cell count in HFR. **b,** vWF staining shows increased number of capillaries and ECs in HFR tissue. EC-Endothelial Cell, HFR-Heart Failure Recovery, vWF-von Willebrand Factor.

**Supplemental Figure 2.**
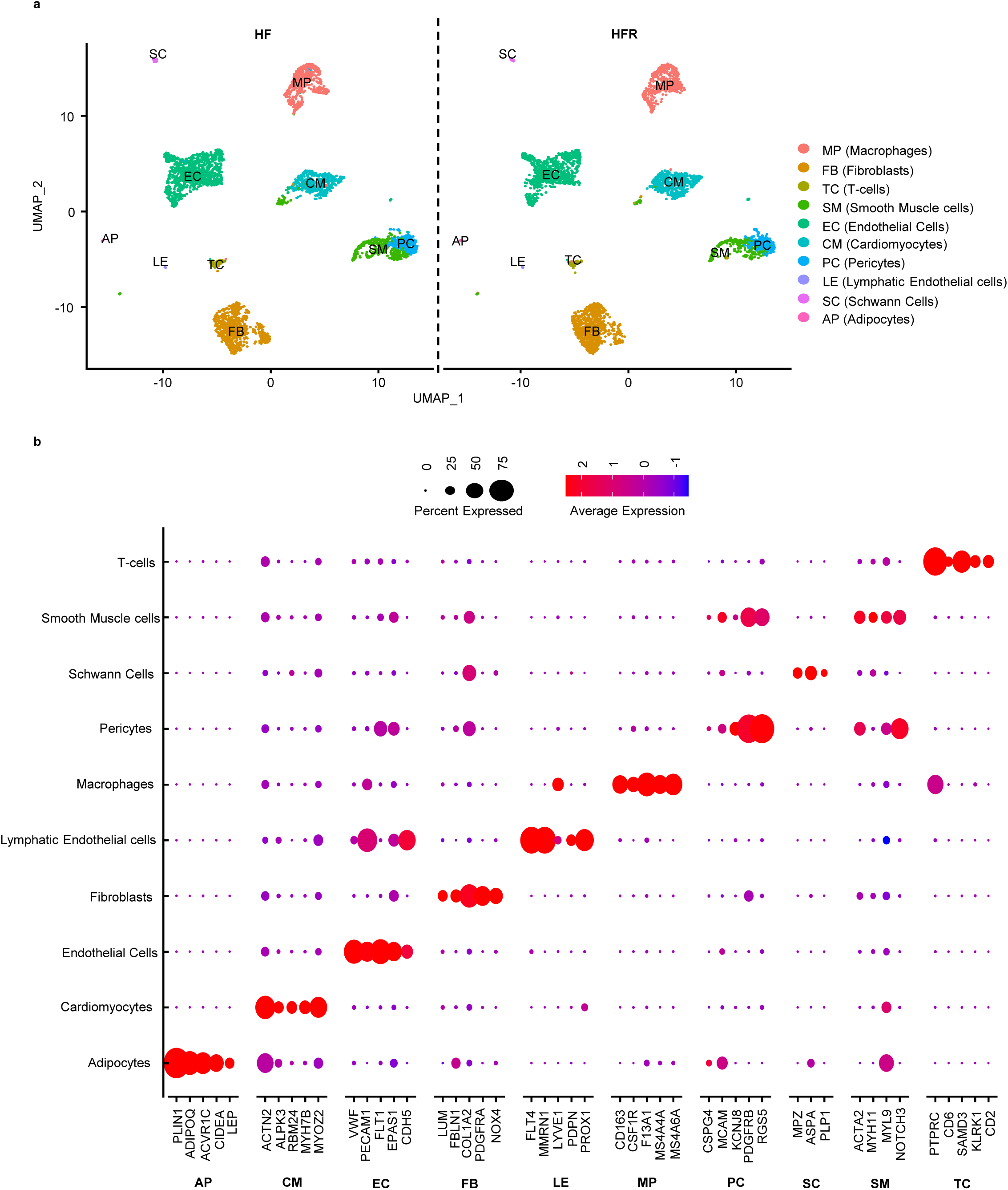
Comprehensive cell type-specific gene expression profiling. **a**, A 2D visualization of the diverse cell subpopulations within heart tissue, identified using Uniform Manifold Approximation and Projection (UMAP) with precise cell type annotations. **b,** A detailed dot plot illustrating the expression patterns of specific gene markers associated with each identified cell type.

**Supplemental Figure 3.**
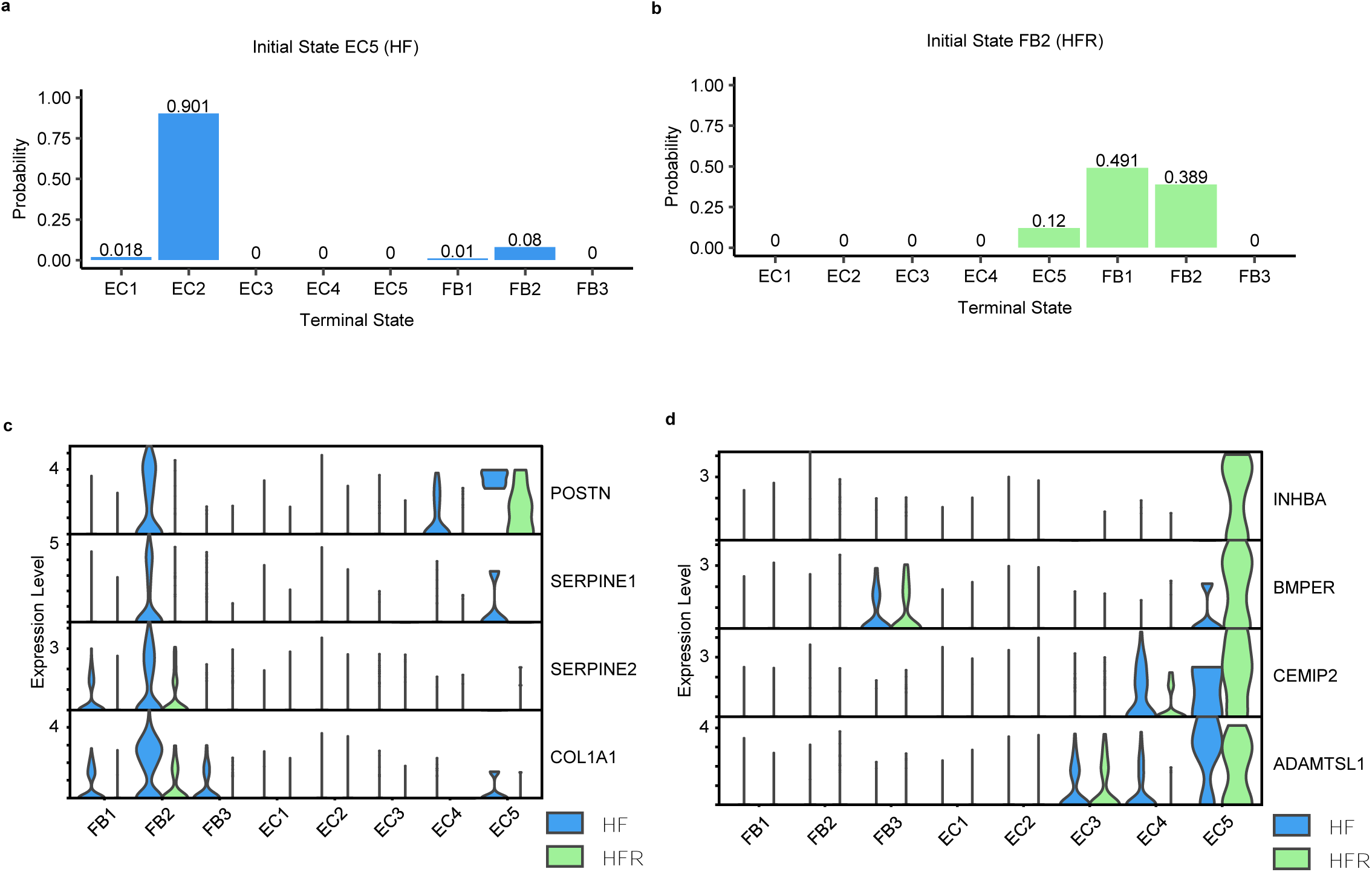
A subtype of fibroblast and endothelial cell corresponds to fibrosis in HF and angiogenesis in HFR, respectively. **a,** RNA velocity rank analysis suggests a transition probability from EC5 to FB2 in HF. **b,** Conversely, RNA velocity rank analysis show transition probability from FB2 to EC5 in HFR. **c,** Fibrosis and tissue repair markers show upregulated expression of FB2 in HF; **d,** Angiogenesis and vascular development related markers are notably upregulated in EC5 of HFR. EC-Endothelial Cell; FB-Fibroblasts; HF-Heart Failure; HFR-Heart Failure Recovery; LVAD-Left ventricular assist device.

**Supplemental Figure 4.**
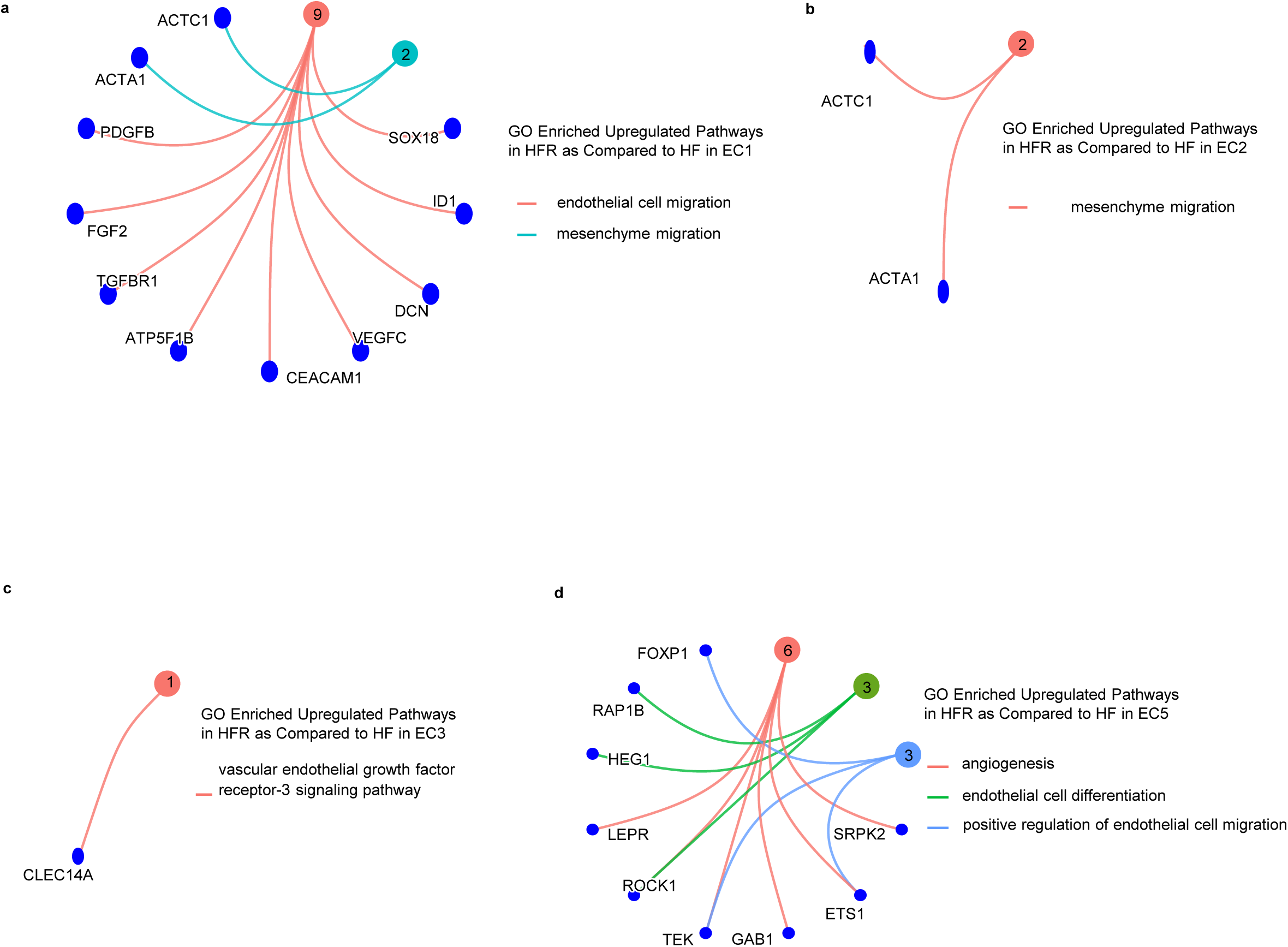
Upregulated angiogenic and vascular pathways in endothelial cells of HFR. **a-d,** CNET plots highlight the specific GO pathway enrichments related to genes upregulated in HFR in endothelial cell subtypes. GO-Gene Ontology, HFR-Heart Failure Recovery. Note-The number of circles denotes the number of upregulated genes in HFR associated with the corresponding pathways.

**Supplemental Figure 5.**
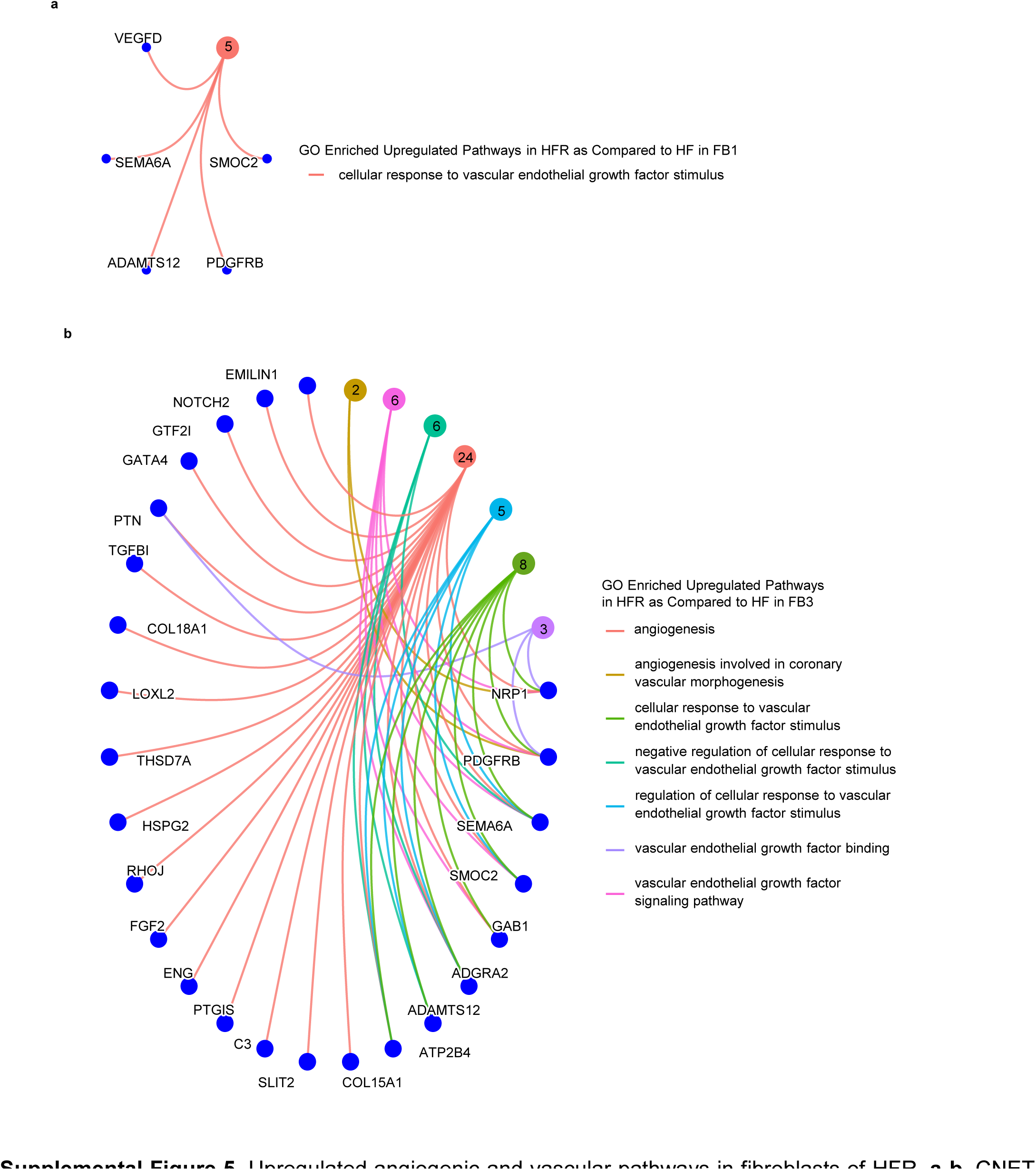
Upregulated angiogenic and vascular pathways in fibroblasts of HFR. **a-b,** CNET plots highlight the specific GO pathway enrichments related to genes upregulated in HFR within the FB subpopulations. FB-Fibroblasts, GO-Gene Ontology, HFR-Heart Failure Recovery. Note-The number of circles denotes the number of upregulated genes in HFR associated with the corresponding pathways.

**Supplemental Figure 6.**
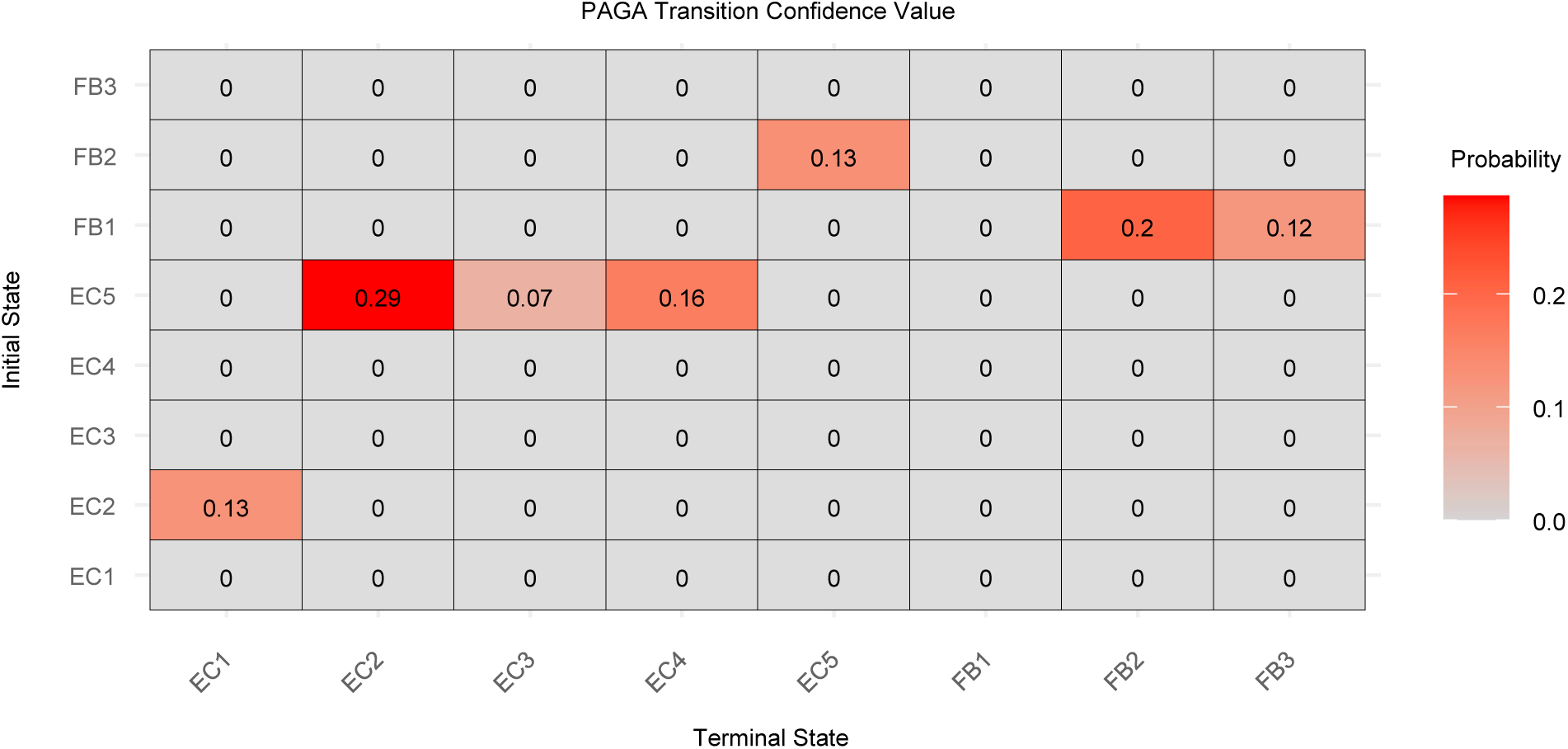
Heatmap shows transition confidence score from initial to terminal states for each pair of subtype of ECs and FBs in HFR. EC-Endothelial cell, FB-Fibroblast, HFR-Heart Failure Recovery, PAGA-Partition based Graph Abstraction.

**Supplemental Figure 7.**
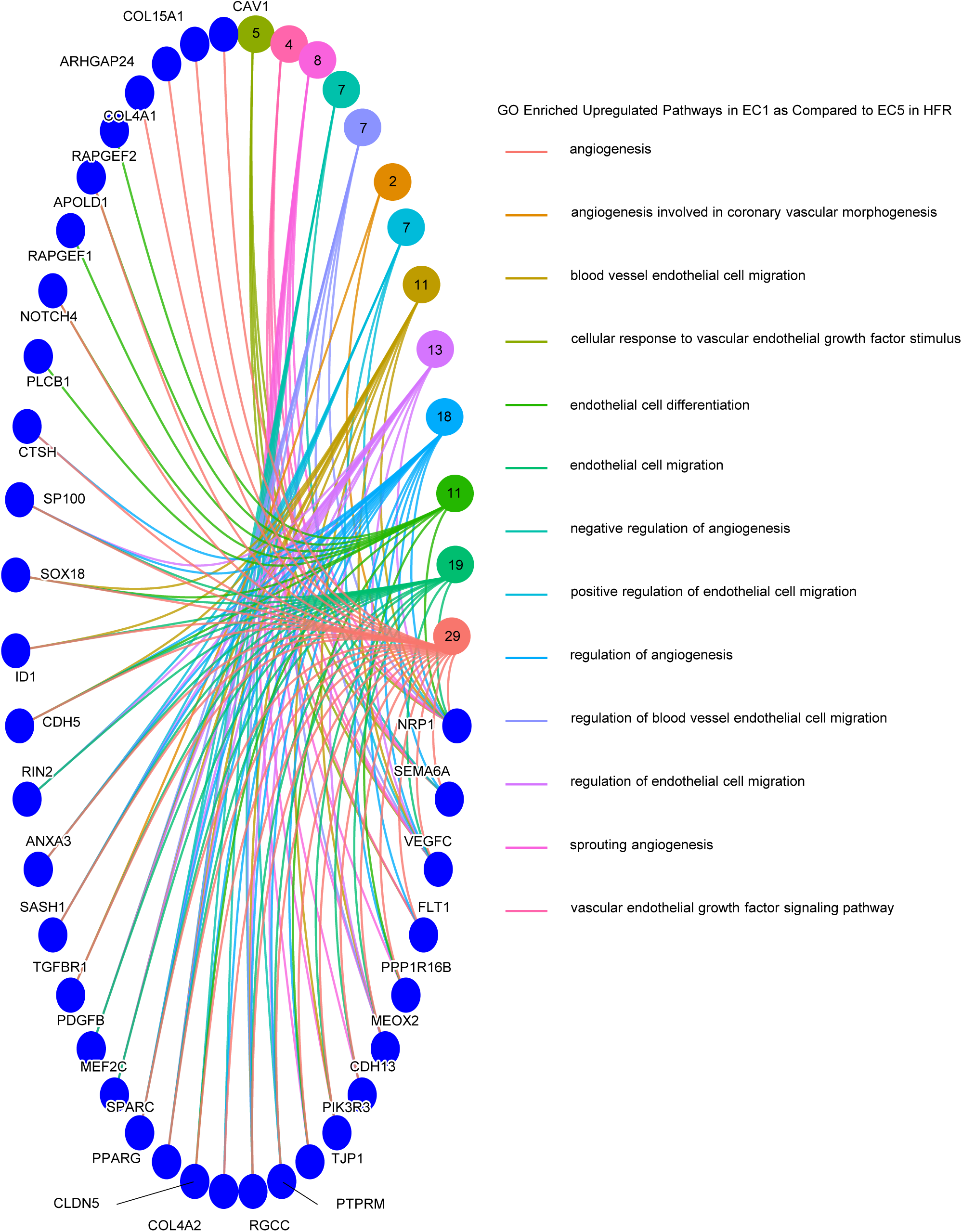
Angiogenic pathways are upregulated in EC1 compared to EC5 in HFR. CNET plot highlighting the specific GO pathway enrichments related to genes upregulated in EC1 compared to EC5 in HFR. EC-Endothelial Cell, GO-Gene Ontology, HFR-Heart Failure Recovery. Note-The number of circles denotes the number of upregulated genes in HFR associated with the corresponding pathways.

**Supplemental Figure 8.**
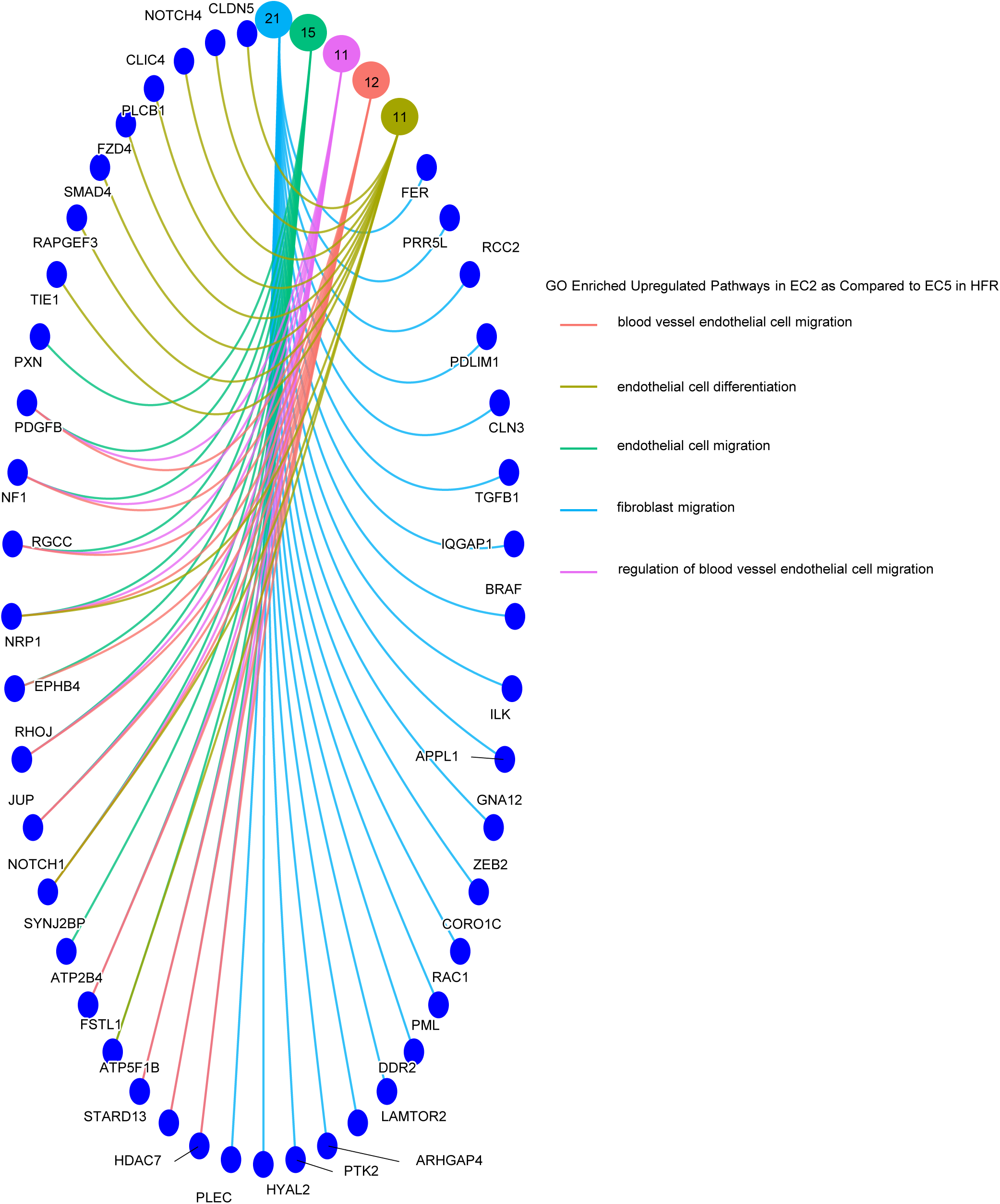
Angiogenic pathways are upregulated in EC2 compared to EC5 in HFR. CNET plot highlighting the specific GO pathway enrichments related to genes upregulated in EC2 compared to EC5. EC-Endothelial Cell, GO-Gene Ontology, HFR-Heart Failure Recovery. Note-The number of circles denotes the number of upregulated genes in HFR associated with the corresponding pathways.

**Supplemental Figure 9.**
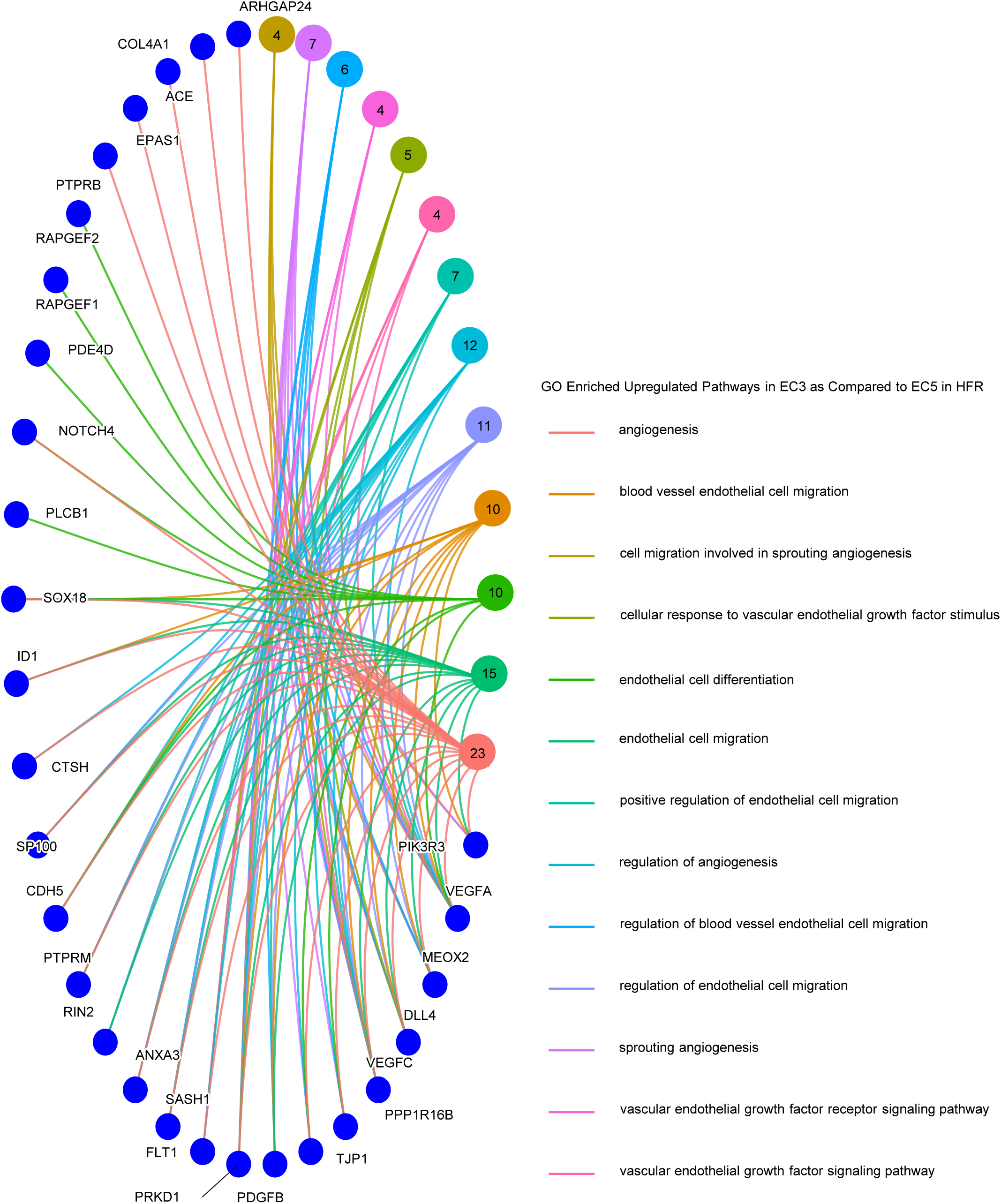
Angiogenic pathways are upregulated in EC3 compared to EC5 in HFR. CNET plot highlighting the specific GO pathway enrichments related to genes upregulated in EC3 compared to EC5. EC-Endothelial Cell, GO-Gene Ontology, HFR-Heart Failure Recovery. Note-The number of circles denotes the number of upregulated genes in HFR associated with the corresponding pathways.

**Supplemental Figure 10.**
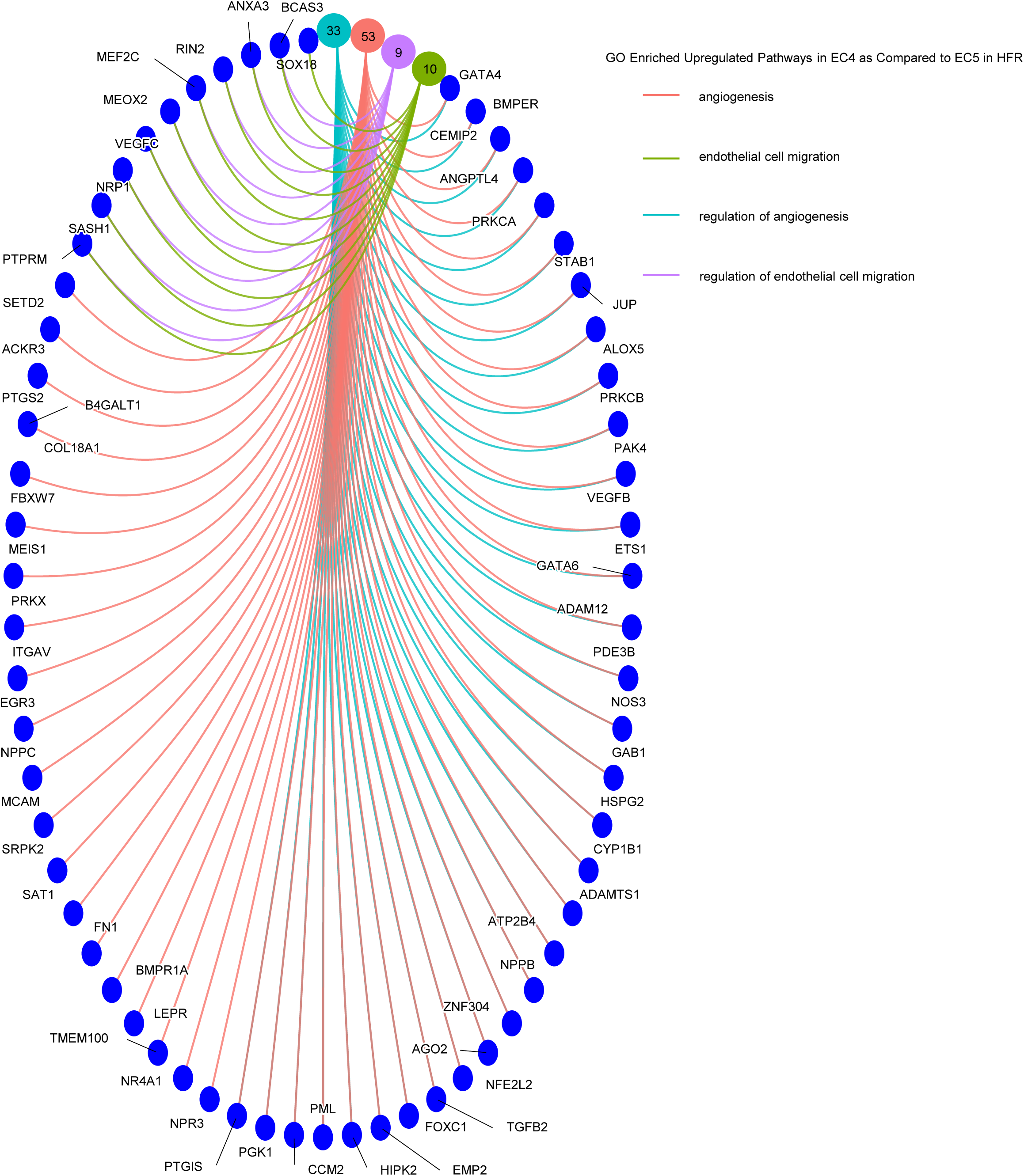
Angiogenic pathways are upregulated in EC4 compared to EC5 in HFR. CNET plot highlighting the specific GO pathway enrichments related to genes upregulated in EC4 compared to EC5. EC-Endothelial Cell, GO-Gene Ontology, HFR-Heart Failure Recovery. Note-The number of circles denotes the number of upregulated genes in HFR associated with the corresponding pathways.

**Supplemental Figure 11.**
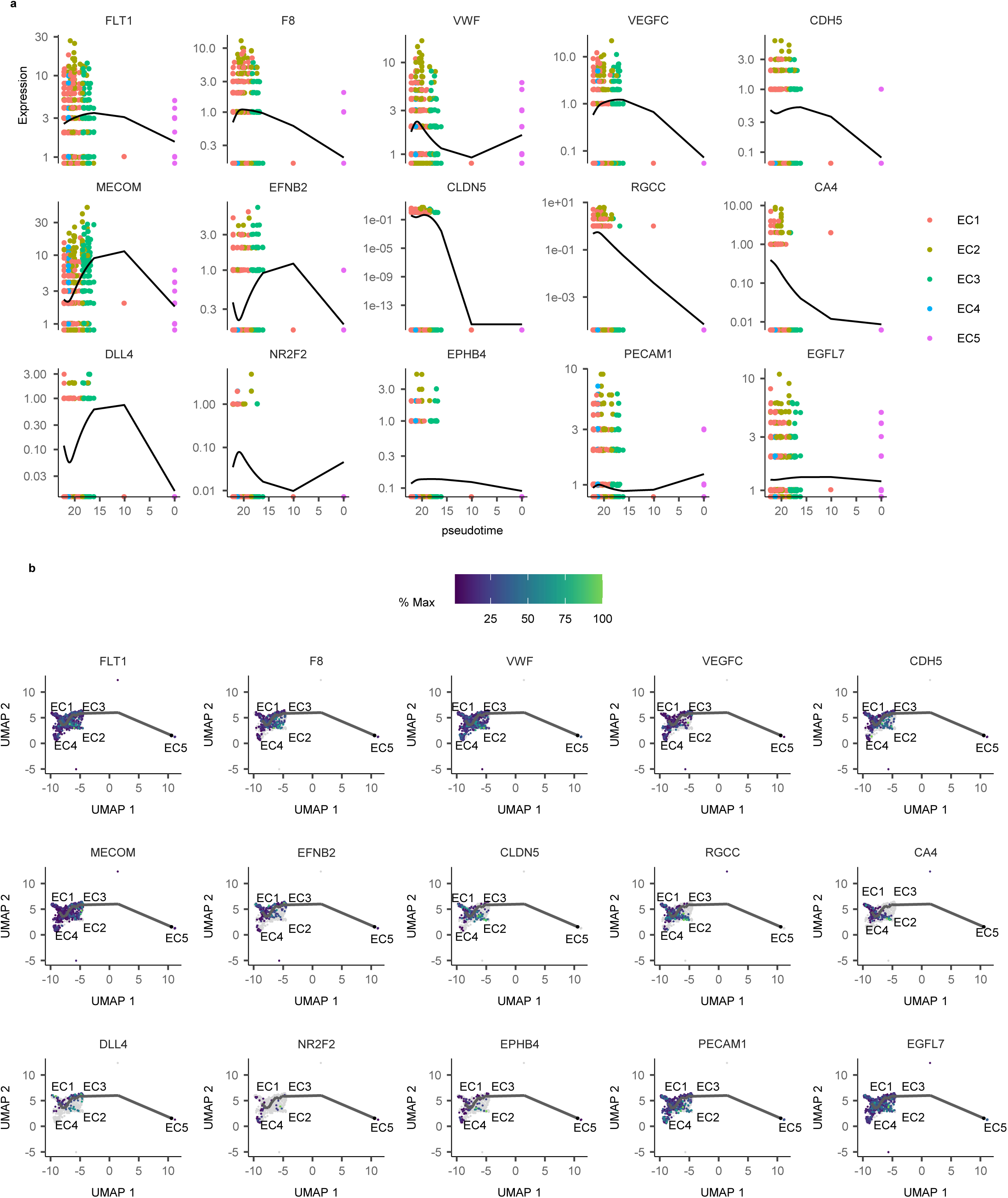
Endothelial cell markers identified in HFR. **a,** Gene expression increases with pseudotime, revealing the transition from EC5 to other endothelial cell subtypes. **b,** Pseudotime percentage indicating the transition dynamics from EC5 to other endothelial cell subtypes.

**Supplemental Figure 12.**
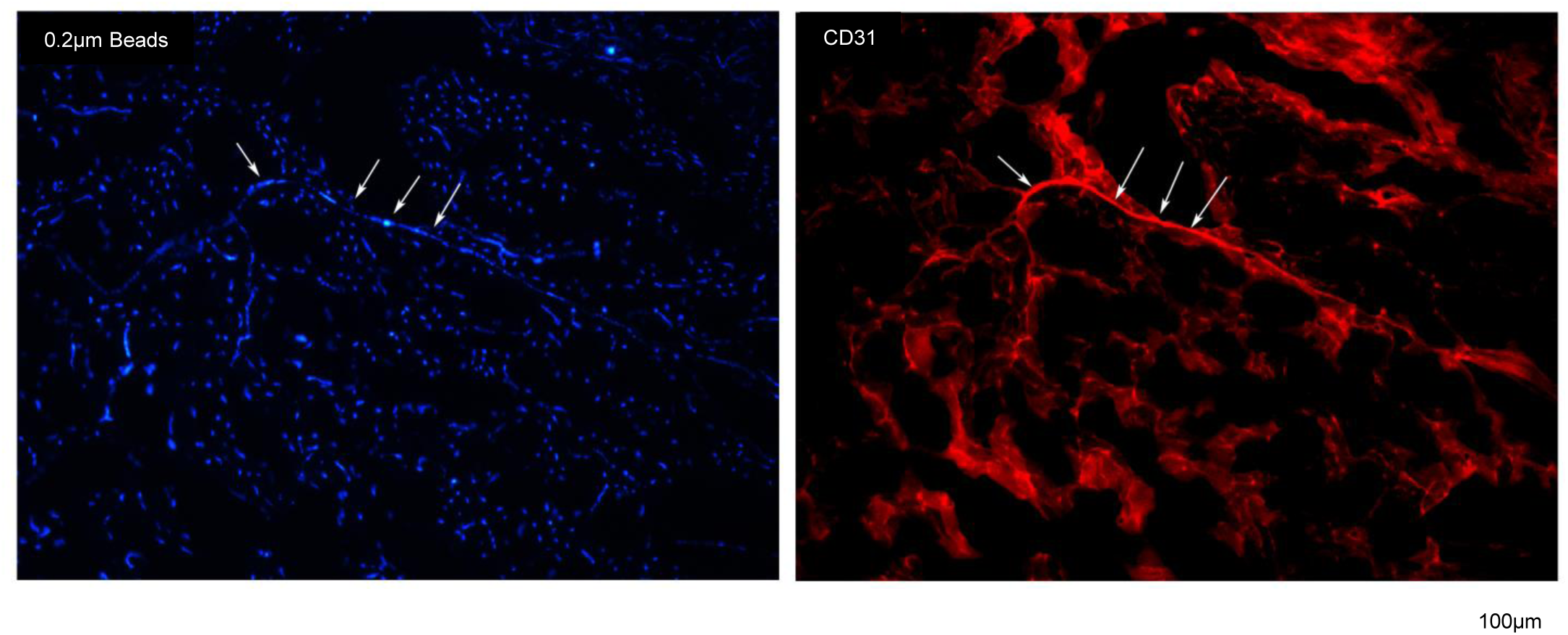
CD31 co-staining with fluorescent microsphere beads. The microsphere beads (blue) and CD31 (red) staining are co-localized (white arrows) suggesting that beads fill up the capillaries and are not leaking out of the vessels.

**Supplemental Figure 13.**
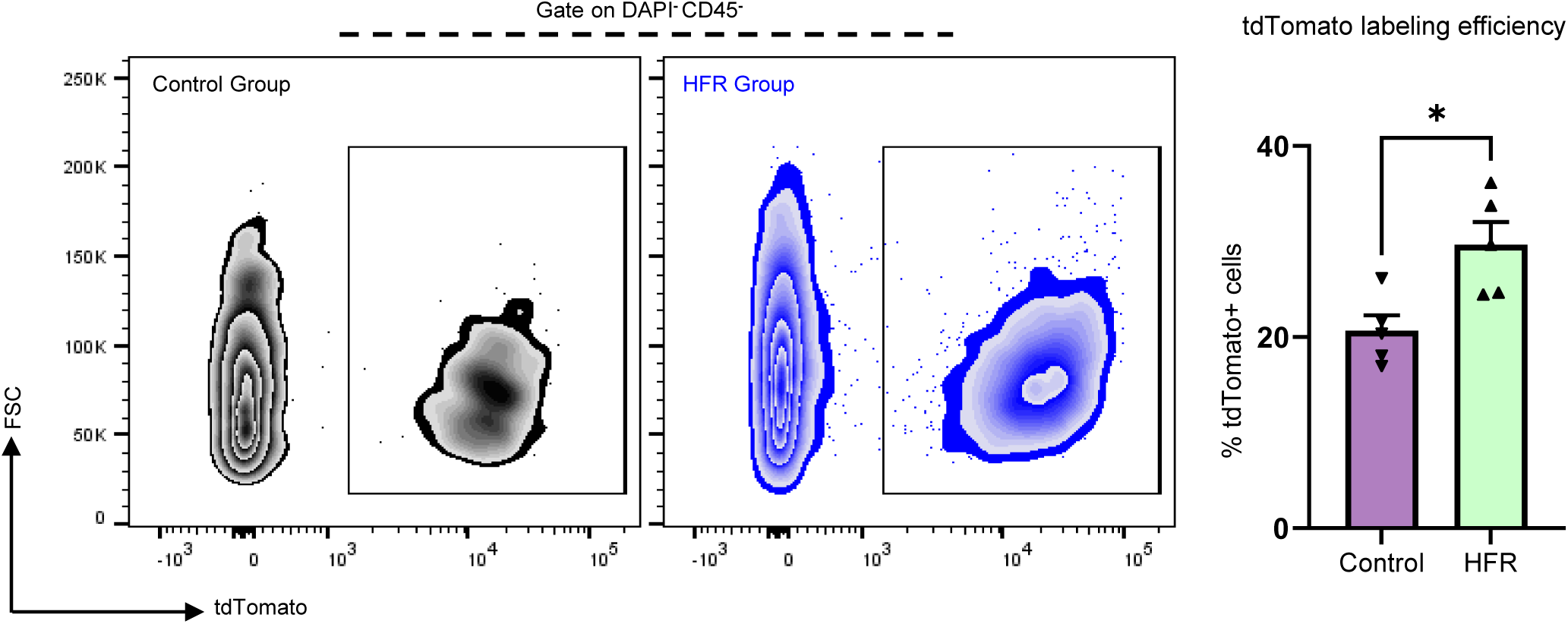
Fibroblast labeling efficiency determined by flow cytometry. The representative plots show the percentages of tdTomato+ cells among live (DAPI-) non-immune cells (CD45-) in both control group and HFR group.

**Supplemental Figure 14.**
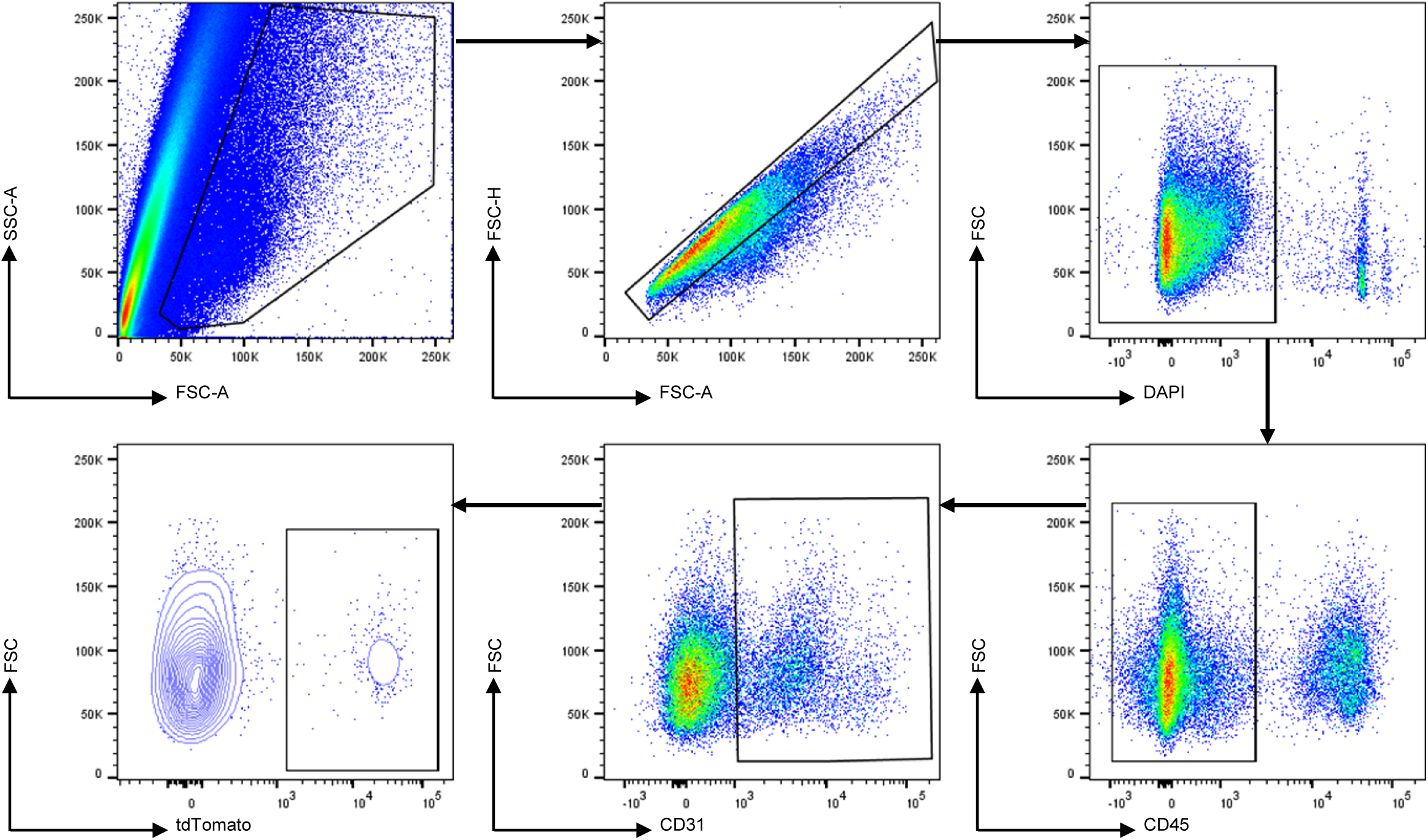
Gating strategy for tdTomato+ cells from CD31+ cells for flow cytometry. Live cells were gated based on DAPI and CD45 positive immune cells which were excluded. CD31 positive endothelial cells were selected and stained for tdTomato and the positive cells were deemed as the transdifferentiation population.

**Supplemental Video.**
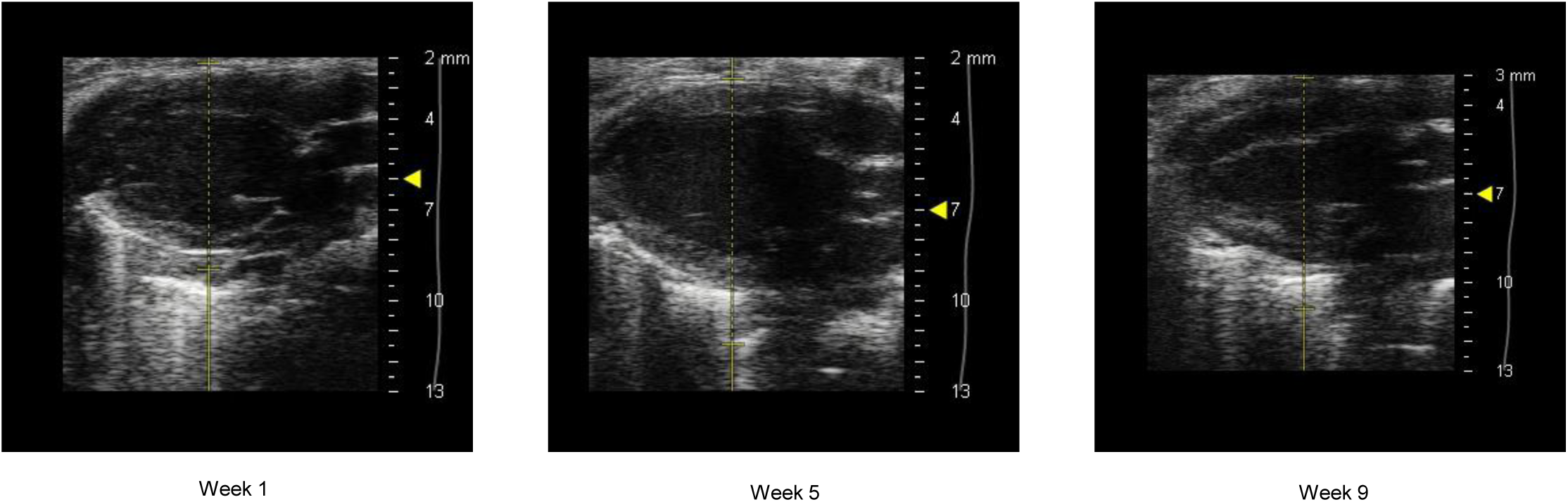
Echocardiography at week 1, week 5 and week 9 of the heart failure and recovery protocol.

## Notes

### Competing Interest Statement

The authors have declared no competing interest.

### Clinical Trial

N/A. This study is not a clinical trial.

### Author Declarations

Cardiac tissue samples were acquired based on an institutional protocol approved by the Houston Methodist Hospital Institutional Review Board [IRB Pro 00006097]. All animal experiments were conducted with approval from the Houston Methodist Research Institute Institutional Animal Care and Use Committee (Houston, TX).

